# miR-151a-5p in Neuron-Derived Extracellular Vesicles Mediates Antidepressant Response

**DOI:** 10.64898/2026.08.11.26360193

**Authors:** Dariusz Żurawek, Alice Morgunova, Laura M. Fiori, Jennie Yang, Reine Khoury, Claudia Belliveau, Maria Antonietta Davoli, Pascal Ibrahim, Anjali Chawla, Sierra Codeluppi-Arrowsmith, Faranak Farzan, Sidney H. Kennedy, Raymond W. Lam, Roumen Milev, Daniel J. Müller, Claudio N. Soares, Suzan Rotzinger, Valerie H. Taylor, Rudolf Uher, Jane A. Foster, Benicio N. Frey, Naguib Mechawar, Cecilia Flores, Corina Nagy, Gustavo Turecki

## Abstract

Major depressive disorder (MDD) lacks accessible molecular markers that reflect brain pathology and monitor treatment response. Using the neuron-specific protein SNAP25, we investigated the cargo of neuron-derived extracellular vesicles (NEVs) isolated from plasma in relation to antidepressant treatment and identified miR-151a-5p as a mediator of treatment response, increasing selectively in responders while remaining low in non-responders. Plasma levels mirrored deficits in human post-mortem brain tissue from the ventral anterior cingulate cortex, a cortical area implicated in MDD. In mice, engineered NEVs enriched with miR-151a-5p delivered cargo selectively to neurons, where miR-151a-5p engaged the RNA-induced silencing complex and regulated genes involved in synaptic networks, including RIMS3 and ELAVL3. Moreover, administration of miR-151a-5p–loaded NEVs in a model of depressive-like behavior produced rapid antidepressant-like effects. Together, these findings identify a vesicle-based mechanism linking peripheral biomarkers to central pathophysiology and demonstrate that miR-151a-5p functions both as a predictor and effector of antidepressant response.

## Introduction

Major depressive disorder (MDD) is a prevalent psychiatric disorder associated with substantial personal, economic, and societal burden, as well as increased mortality^1^. Although existing medications alleviate symptoms in many patients, approximately 30–40% fail to respond to first-line treatment and undergo prolonged trial-and-error regimens^2^. Even among responders, relapse rates approach 59% after therapy discontinuation^3^, underscoring the inadequate sustainability of current strategies. Unfortunately, therapeutic innovation is currently limited by insufficient understanding of the molecular alterations underlying both MDD and antidepressant mechanisms of action.

Peripheral samples do not directly reflect brain molecular processes, and sampling brain tissue of living individuals is not feasible in most clinical contexts, necessitating the identification of accessible markers of brain-relevant biology. In this context, extracellular vesicles (EVs), which are nanoscale membrane-bound vesicles released by all cells, including neurons and glial cells^4,5^, offer a promising solution. EVs contain molecular cargo, such as proteins^6–8^, lipids^6,7^, and diverse RNA species^8,9^, which reflect the composition of their cell of origin^9^. Moreover, EVs carry surface protein markers that mediate selective uptake by recipient cells and can be used to infer their origin^10,11^. Beyond their value as potential biomarkers, EVs play an active role in intercellular communication. Neurons, in particular, use EV-mediated signaling to exchange molecular information outside of classical synaptic transmission, adding an extrasynaptic layer of communication^12–14^. EVs protect their cargo from enzymatic degradation and circulate systemically, enabling detection in peripheral biofluids^15^.

Among EV cargo molecules, microRNAs (miRNAs) are particularly interesting, as they regulate gene expression post-transcriptionally and enable coordinated regulation of multiple molecular networks. The brain is rich in tissue-specific miRNAs^16,17^ and neurons are especially enriched in miRNAs involved in synaptic function and plasticity^17,18^. Converging evidence implicates miRNAs in the pathophysiology of psychiatric disorders^19–21^, and supports their therapeutic potential in neuropsychiatric disorders^22^.

Here, we identified SNAP25 as a neuron-specific marker, enabling the isolation of neuron-derived extracellular vesicles (NEVs) from human plasma and brain tissue with comparable molecular compositions. NEV miRNA profiling identified treatment-dependent regulation of miR-151a-5p that distinguished antidepressant responders from non-responders. Notably, pre-treatment plasma NEV miR-151a-5p levels in individuals with MDD mirrored those observed in NEVs isolated from post-mortem ventral anterior cingulate cortex (vACC) of individuals who died during an episode of MDD. Finally, using engineered NEVs (eNEVs) enriched with miR-151a-5p, we demonstrated neuron-selective delivery in mice and showed that miR-151a-5p transfer modulated neuronal function and induced antidepressant-like behavioral effects in a mouse model of chronic social defeat stress.

## Results

### 1. NEVs from brain and plasma are SNAP25-positive and carry neuron-specific markers

To identify NEVs in human brain and plasma, we focused on the synaptic protein SNAP25 as a candidate marker of neuronal origin. Because the field lacks a robust marker specific to neurons and NEVs, we analyzed GTEx data to compare tissue-specific mRNA and protein levels of SNAP25 with other candidates proposed in the literature^23–27^. We found that SNAP25 showed the highest brain enrichment and specificity, whereas alternative markers displayed broader tissue distribution (Fig. 1A). Next, we asked whether this specificity extended across neuronal populations. Analysis of single-cell transcriptomic data from the human prefrontal cortex (PFC) revealed robust SNAP25 expression across multiple neuronal subtypes^28^, while other candidates showed either low neuronal expression or broader, non-neuronal profiles (Fig. 1B). To determine whether SNAP25 associates with EVs in brain tissue and biofluids, we performed size-exclusion chromatography (SEC) followed by enzyme-linked immunosorbent assays (ELISAs) on fresh human brain, cerebrospinal fluid (CSF), and plasma. We found that SNAP25 co-eluted with canonical EV markers (CD9, CD63, CD81) in EV-rich fractions and was absent from later fractions containing free protein (Fig. 1C), indicating that SNAP25 is vesicle-associated rather than soluble in all sample types. As controls for soluble protein, we assessed endogenous levels of albumin, as well as recombinant SNAP25 dissolved in buffer and subjected to SEC–ELISA, and both controls were found to be eluted in non-vesicular fractions (Fig. 1C). We next examined whether SNAP25 defines a distinct EV subpopulation using transmission electron microscopy (TEM) with immunogold co-labeling for pan-tetraspanins (CD9, CD63, CD81) and SNAP25. We found EVs from human cortex and plasma that simultaneously displayed both markers on their surface, directly visualizing SNAP25-positive vesicles and supporting their neuronal origin (Fig. 1D). Guided by these observations, we selectively isolated SNAP25-positive EVs by immunocapture. These vesicles exhibited the characteristic EV morphology and a size distribution centered around ∼100 nm, confirming their structural integrity (Fig. 1E). Molecular profiling further supported their central neuronal identity: SNAP25-immunocaptured EVs were enriched in neuronal (NCAM1) and canonical EV markers (Alix, CD9), while lacking markers of peripheral neurons (PRPH), glia (GFAP), intracellular compartments (CANX), and lipoproteins (ApoB), arguing against contamination and indicating a predominantly central neuronal origin (Fig. 1F). Finally, to define SNAP25 topology around the EV membrane, we treated intact and detergent-permeabilized EVs with Botulinum neurotoxin A light chain (BoNT/A), which selectively degrades SNAP25 (Fig. 1G). BoNT/A partially reduced SNAP25 levels in intact brain (Fig. 1H) and plasma (Fig. 1I) EVs, whereas detergent permeabilization led to complete signal loss, suggesting SNAP25 is localized on both the external and luminal surfaces of NEV membranes. Together, these data established SNAP25 as a specific, surface-accessible marker of NEVs and validated its use for their selective isolation from both brain and plasma.

**Fig. 1:**
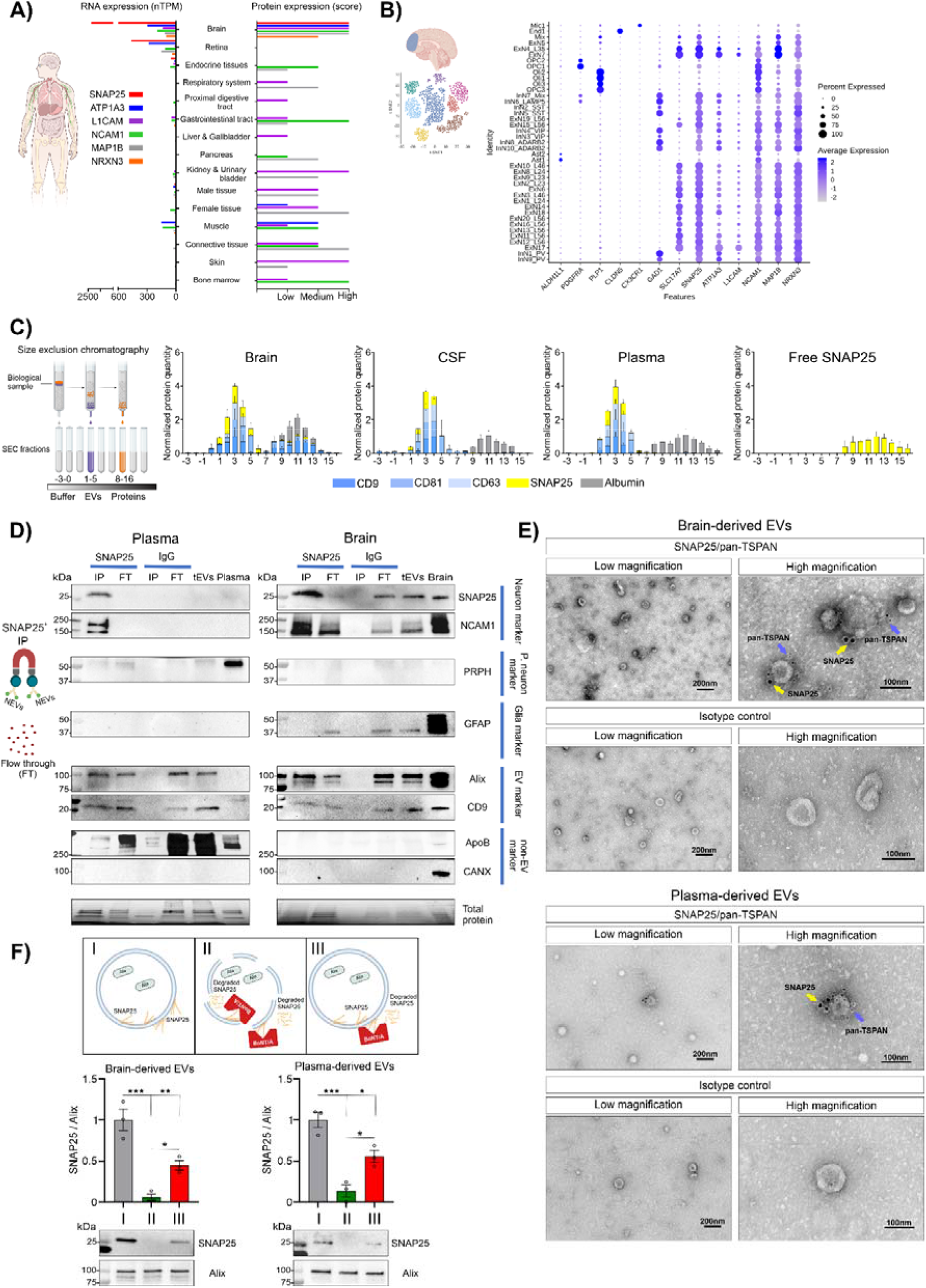
SNAP25 identifies neuron-derived extracellular vesicles (NEVs) in human brain and plasma. A) Comparison of transcript abundance (nTPM) and protein expression scores for SNAP25 and other candidate NEV markers (ATP1A3, L1CAM, NCAM1, MAP1B and NRXN3) across human tissues. SNAP25 showed the strongest enrichment in brain tissue at both the RNA and protein levels, with no detectable expression in peripheral tissues, whereas other candidate markers showed lower brain specificity or broader tissue expression. RNA expression data were obtained from GTEx, and protein expression scores were obtained from the Human Protein Atlas. B) Single-nucleus RNA-seq analysis of SNAP25 and candidate NEV markers expression across cell populations in the human dorsolateral prefrontal cortex (dlPFC). Dot size indicates the percentage of cells expressing each gene, and color denotes average scaled expression. SNAP25 was broadly and selectively expressed across neuronal subpopulations, whereas other candidate markers showed lower specificity or were absent from selected neuronal populations. Data were derived from previously published publicly available human dlPFC snRNA-seq datasets generated by our laboratory. C) ELISA-based analysis of SNAP25, albumin and canonical EV markers CD9, CD81 and CD63 across SEC fractions from human brain tissue (n = 3), CSF (n = 3), plasma (n = 3) and free SNAP25 diluted in PBS (n = 3). SNAP25 was detected in EV-enriched SEC fractions, where it co-eluted with canonical EV markers in brain, CSF and plasma. Albumin was detected only in late SEC fractions corresponding to soluble proteins, as was free SNAP25 in PBS. In brain preparations, two SNAP25 peaks were observed, corresponding to EV-enriched fractions and later fractions containing soluble proteins and very small particles. For visualization, data were normalized independently for each target within each tissue or biofluid across all SEC fractions, because absolute concentration ranges differed substantially between proteins. For each target, the SEC fraction with the highest measured concentration was assigned a value of 1, and all other fractions are shown as relative values proportional to that maximum signal. D) Representative TEM images showing immunogold labeling of total EV populations isolated from human brain (n = 3; upper panels) and plasma (n = 3; lower panels). EVs were labeled for SNAP25 using 12-nm gold particles (yellow arrows) and pan-tetraspanins (CD9, CD81 and CD63) using 6-nm gold particles (blue arrows). Isotype control antibodies were used to assess labeling specificity. Low-magnification images show overall EV labeling patterns (scale bar, 200 nm), and high-magnification images show EVs with co-localized SNAP25 and pan-tetraspanin signal on their surface (scale bar, 100 nm). E) Characterization of SNAP25-based immunocapture of NEVs from human brain (n = 3) and plasma (n = 3). TEM images (upper panels) show that SNAP25-immunocaptured NEVs from brain and plasma retained typical vesicular morphology. Nanoparticle tracking analysis (NTA; lower panels) showed a particle size distribution peaking at approximately 100 nm in both NEVs from brain (n=3) and plasma (n=3), consistent with the expected EV size range. F) Western blot characterization of SNAP25-immunocaptured NEVs from plasma (n = 3) and brain (n = 3). NEVs from both sources were enriched in SNAP25 and the neuronal marker NCAM1, but lacked PRPH, a marker of peripheral neurons, GFAP, a glial marker, and the non-EV markers ApoB and CANX. NEVs from brain and plasma also contained the canonical EV markers ALIX and CD9. The enrichment of SNAP25 and NCAM1 together with the absence of PRPH supports a predominantly central, rather than peripheral, neuronal origin of these vesicles. IP - SNAP25-immunocaptured NEVs; FT - flow-through after immunocapture; tEVs - total EV population; PL - total plasma protein; BR - total brain protein; SNAP25 - immunocapture with anti-SNAP25 antibody; IgG - immunocapture with isotype control antibody; P.neuron marker – Peripheral neuron marker. G) Schematic of the BoNT/A protection assay used to determine SNAP25 topology in EVs. EVs were analyzed under three conditions: (i) untreated EVs, used as controls; (ii) EVs incubated with BoNT/A (2.5 μg) and Triton X-100 (1%), which disrupts EV membranes and permits enzymatic access to both luminal and externally exposed SNAP25; and (iii) EVs incubated with BoNT/A alone, in which intact membranes protect luminal SNAP25 while allowing digestion of externally exposed SNAP25. H) Representative western blots (lower panel) and quantification (upper panel) of SNAP25 levels normalized to ALIX in brain EVs after BoNT/A protection assay. BoNT/A reduced SNAP25 levels in intact EVs, whereas combined BoNT/A and Triton X-100 treatment produced a stronger loss of SNAP25 signal, consistent with SNAP25 localization in both externally accessible and protected intravesicular compartments. One-way ANOVA: F (2) = 31.05, P = 0.0007; Tukey’s post hoc test: i vs ii, P < 0.001; i vs iii, P < 0.01; ii vs iii, P < 0.05; n = 3 per group. I) Representative western blots (lower panel) and quantification (upper panel) of SNAP25 levels normalized to ALIX in plasma EVs after BoNT/A protection assay. BoNT/A reduced SNAP25 levels in intact EVs, whereas combined BoNT/A and Triton X-100 treatment produced a stronger loss of SNAP25 signal, consistent with SNAP25 localization in both externally accessible and protected intravesicular compartments. One-way ANOVA: F (2) = 29.50, P = 0.0008; Tukey’s post hoc test: i vs ii, P < 0.001; i vs iii, P < 0.01; ii vs iii, P < 0.05; n = 3 per group. For all graphs, data are presented as mean ± s.e.m.

### 2. miR-151a-5p levels in NEVs from plasma of individuals with depression reflect MDD-related alterations in the brain and are markers of antidepressant treatment response

To test whether the molecular cargo of NEVs reflects MDD-related changes and antidepressant treatment response, we analyzed plasma samples from 142 adult individuals with MDD (89 females, 53 males) and 73 psychiatrically healthy controls (48 females, 25 males) who participated in the Canadian Biomarker Integration Network in Depression (CAN-BIND-1, <u>NCT01655706</u>) clinical trial of antidepressant response^29^. Briefly, individuals with MDD received eight weeks of the selective serotonin reuptake inhibitor escitalopram (ESC) treatment, with symptom severity assessed at baseline (W0) and after treatment (W8) using the Montgomery–Åsberg Depression Rating Scale (MADRS), while healthy controls received no pharmacological intervention and were evaluated at matched time points (Fig. 2A). All groups were matched for demographic variables (Supplementary Fig. 1A). Post-treatment assessments, based on established clinical criteria^29^, were used to classify patients as responders or non-responders (Supplementary Fig. 1B). Importantly, responders and non-responders exhibited indistinguishable plasma levels of ESC, its major metabolite S-desmethylcitalopram (S-DCT) (Supplementary Fig. 1C), and the ratio between both (Supplementary Fig. 1D), throughout treatment, indicating that clinical differences in response to treatment were not attributable to drug exposure or its pharmacokinetics, but rather to other mechanisms. We next isolated NEVs from plasma collected at W0 and W8 and profiled their miRNA cargo via RNA-sequencing (Fig. 2A). Of the miRNAs detected in NEVs, we identified a significant interaction between treatment response and time for miR-151a-5p (GLZ: Wald: W (2) = 13.43, P = 0.0012, FDR = 0.0351) indicating that its levels change over time as a function of clinical outcome (Supplementary Fig. 1E). Both sequencing (Fig. 2B and D) and RT–qPCR validation (Fig. 2C and E) confirmed that miR-151a-5p levels increased from W0 to W8 only in responders (RNA-seq log2 FC = 0.71, P = 0.05; RT-PCR log2 FC = 0.80, P < 0.01) reaching levels comparable to healthy controls, which displayed stable expression over time, whereas non-responders showed persistently low expression over time, and compared to other groups (RNA-seq log2 FC = - 0.24, P = 0.05; RT-PCR log2 FC = - 0.36, P < 0.01). Importantly, these changes were not driven by altered vesicle release or peripheral miRNA expression, as total plasma NEV concentrations were similar across groups and time points (Supplementary Fig. 1F-G), and miR-151a-5p levels remained unchanged in leukocytes from the same individuals (Supplementary Fig. 1H-I), indicating that this effect is specific to NEVs.

**Fig. 2:**
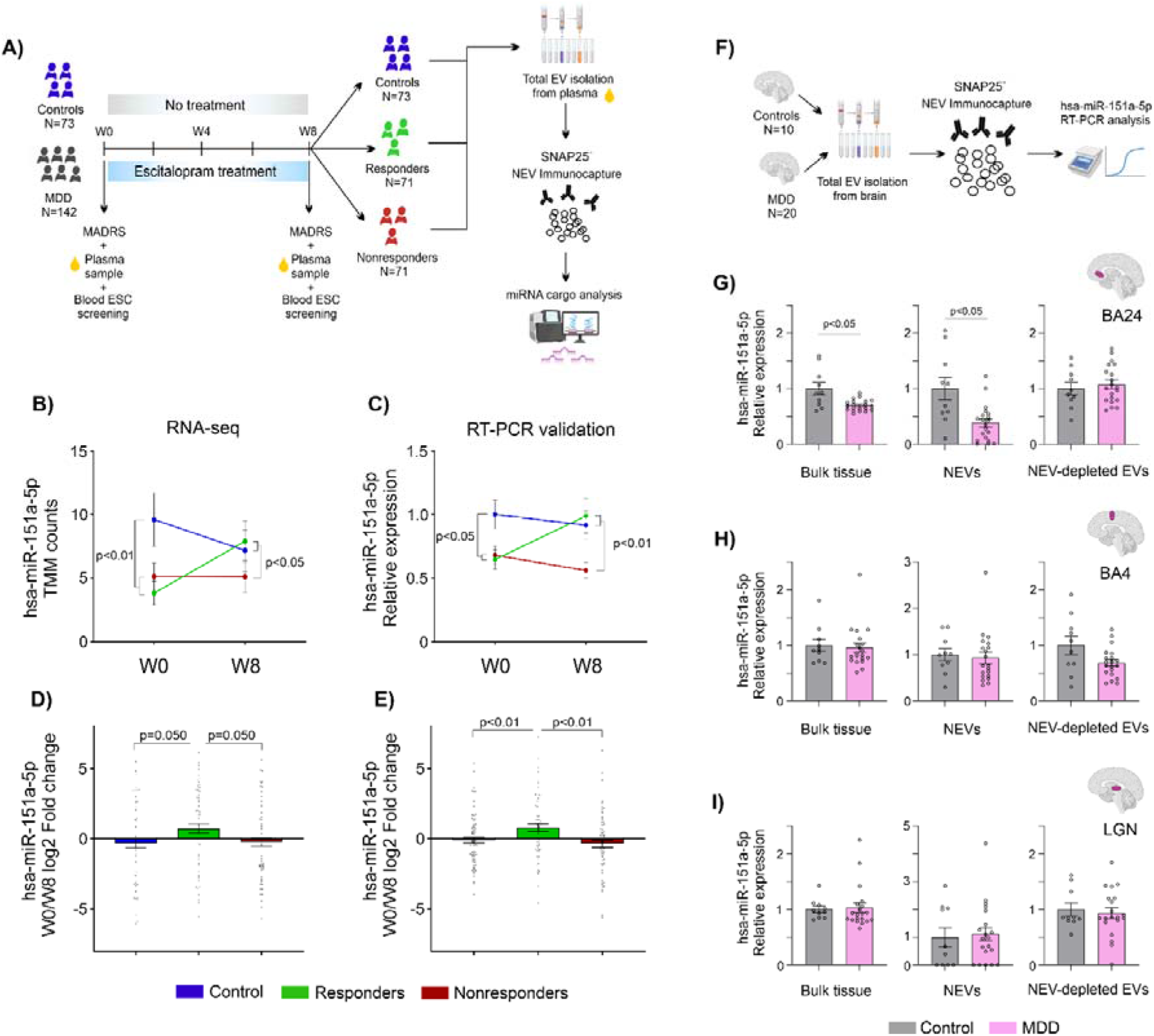
Plasma NEV miR-151a-5p levels reflect MDD-associated brain alterations and serve as a marker of antidepressant treatment response. A) Schematic of the CAN-BIND1 clinical study design. The cohort included healthy control participants (n = 73) and MDD patients (n = 142). MDD patients received ESC treatment for 8 weeks, and depressive symptoms were assessed using the MADRS scale before treatment (week 0, W0) and after treatment (week 8, W8). Based on MADRS score reduction at W8, patients were classified as responders (n = 71) or non-responders (n = 71). Plasma was collected from all participants at both time points, NEVs were isolated, and their miRNA cargo was analyzed by small RNA-seq and later validated by RT-PCR. B) Small RNA-seq analysis of miR-151a-5p levels in plasma NEVs, expressed as TMM-normalized counts, at W0 and W8 in controls, responders and non-responders. Generalized linear model (GLZ) with negative binomial distribution showed a significant interaction between time and treatment response group. Wald: W (2) = 13.43, P = 0.0012, FDR = 0.0351. Before treatment (W0), patients with MDD showed significantly lower miR-151a-5p levels in plasma NEVs than controls, with no difference between future responders and non-responders. GLZ post hoc tests: W0 controls (n = 73) vs W0 responders (n = 71), P < 0.01; W0 controls vs W0 non-responders (n = 71), P < 0.01; W0 responders vs W0 non-responders, P = 0.11. After 8 weeks of ESC treatment (W8), miR-151a-5p levels remained low in non-responders but increased in responders to levels comparable to controls. GLZ post hoc tests: W8 controls (n = 73) vs W8 responders (n = 71), P = 0.43; W8 controls vs W8 non-responders (n = 71), P < 0.05; W8 responders vs W8 non-responders, P < 0.05. C) RT–qPCR validation of miR-151a-5p levels in plasma NEVs, expressed relative to miR-320a-3p, confirmed observations from small RNA-seq. Two-way ANOVA showed a significant time × treatment response interaction: F (2,417) = 3.332, P = 0.0367. Before treatment (W0), patients with MDD showed significantly lower miR-151a-5p levels in plasma NEVs than controls, with no difference between future responders and non-responders. Benjamini, Krieger and Yekutieli two-stage linear step-up post hoc test: W0 controls (n = 72) vs W0 responders (n = 69), P < 0.05; W0 controls vs W0 non-responders (n = 68), P < 0.05; W0 responders vs W0 non-responders, P = 0.28. After 8 weeks of ESC treatment (W8), miR-151a-5p levels remained low in non-responders but increased in responders to levels comparable to controls. Benjamini, Krieger and Yekutieli two-stage linear step-up post hoc test: W8 controls (n = 72) vs W8 responders (n = 71), P = 0.21; W8 controls vs W8 non-responders (n = 71), P < 0.01; W8 responders vs W8 non-responders (n = 71), P < 0.01. D) Log2 fold change in plasma NEV miR-151a-5p levels between W0 and W8 in controls, responders and non-responders, measured by small RNA-seq. An increasing trend in miR-151a-5p levels over time was observed only in responders. One-way ANOVA: F (2) = 3.39, P = 0.036; Benjamini, Krieger and Yekutieli two-stage linear step-up post hoc test: controls (n = 73) vs responders (n = 71), P = 0.053; controls vs non-responders (n = 71), P = 0.879; responders vs non-responders, P = 0.053. E) RT–qPCR validation of log2 fold change in plasma NEV miR-151a-5p levels between W0 and W8 confirmed a significant increase in responders. One-way ANOVA: F (2) = 5.884, P = 0.0033; Benjamini, Krieger and Yekutieli two-stage linear step-up post hoc test: controls (n = 71) vs responders (n = 69), P < 0.01; controls (n = 73) vs non-responders (n = 71), P = 0.165; responders (n = 71) vs non-responders (n = 68), P < 0.01. F) Schematic of the post-mortem brain study. Brain tissue from vACC, BA4, LGN was collected from control (n = 10) and MDD (n = 20) subjects. Groups did not differ in racial composition, age, brain pH, PMI, refrigeration delay or RIN. miR-151a-5p levels were measured by RT–qPCR in bulk tissue from all three regions. NEVs were then isolated from remaining tissue by immunoprecipitation, and the flow-through fraction was used as the NEV-depleted EV fraction. miR-151a-5p levels were measured in NEVs and NEV-depleted EVs from each region. G) RT–qPCR analysis of miR-151a-5p levels in vACC. miR-151a-5p was significantly reduced in MDD samples compared with controls in bulk tissue (left panel; two-tailed t-test, P < 0.05) and in NEVs (middle panel; two-tailed t-test, P < 0.05), but not in NEV-depleted EVs (right panel; two-tailed t-test, P = 0.58). H) RT–qPCR analysis of miR-151a-5p levels in BA4. No significant differences were observed between control and MDD samples in bulk tissue (left panel; Mann–Whitney test, P = 0.71), NEVs (middle panel; Mann–Whitney test, P = 0.44) or NEV-depleted EVs (right panel; two-tailed t-test, P = 0.10). I) RT–qPCR analysis of miR-151a-5p levels in LGN. No significant differences were observed between control and MDD samples in bulk tissue (left panel; Mann–Whitney test, P = 0.32), NEVs (middle panel; Mann–Whitney test, P = 0.67) or NEV-depleted EVs (right panel; Mann–Whitney test, P = 0.91). For all graphs, data are presented as mean ± s.e.m.

To determine whether plasma NEV miR-151a-5p reflects MDD brain alterations, we examined post-mortem human brain samples from 10 psychiatrically healthy individuals and 20 individuals with MDD (Fig. 2F and Supplementary Fig. 1J). We isolated NEVs, non-NEVs, and bulk tissue, from three brain regions: the vACC, previously implicated in MDD^30,31^ and antidepressant response^31^, and two control regions not known to be involved in the pathophysiology of MDD, the primary motor cortex (BA4) and the lateral geniculate nucleus (LGN). Only vACC from MDD subjects showed a significant reduction in miR-151a-5p levels in both bulk tissue (two-tailed t-test, t_9.743_ = 2.675, P < 0.05) and NEVs (two-tailed t-test, t_11.43_ = 2.856, P < 0.05) (Fig. 2G), whereas BA4 (Fig. 2H) and LGN (Fig. 2I) displayed similar levels to controls. Together, these findings suggest that SNAP25-enriched NEVs capture brain specific molecular alterations associated with MDD and treatment response, supporting plasma NEV miR-151a-5p as a peripheral marker of MDD and a dynamic indicator of antidepressant efficacy.

### 3. Engineered NEVs enriched with miR-151a-5p selectively deliver cargo to neurons resulting in biologically significant regulation of neuron-enriched genes

To investigate whether NEVs may be involved in intercellular communication and if they actively deliver miR-151a-5p to specific target cells, we produced engineered NEVs with enriched miR-151a-5p cargo (eNEVs). To do so, we isolated native NEVs from fresh mouse cortex and transfected them with fluorescently labeled miR-151a-5p (Supplementary Fig. 2A). Transfection efficiency, purification, and optimal miRNA-to-NEV ratios were verified using multiple independent approaches to maximize vesicular loading while minimizing free miRNA in solution (Supplementary Fig. 2B). TEM and nanoparticle tracking analysis (NTA) showed that eNEVs retained their intact morphology (Supplementary Fig. 2C) and size distribution (Supplementary Fig. 2D). Nano-flow cytometry confirmed efficient miRNA encapsulation: detergent treatment abolished fluorescence, demonstrating that miR-151a-5p was contained within intact vesicles rather than present as extracellular miRNA–reagent aggregates or bound to the EV corona (Supplementary Fig. 2E). Western blotting further showed that eNEVs preserved the protein composition of native NEVs, including SNAP25 and canonical EV markers (Tsg101, Cd9), while lacking markers of cellular contamination (Bip, Vdac) (Supplementary Fig. 2F).

Following stereotaxic injection of eNEVs into the prelimbic and infralimbic regions of mouse medial prefrontal cortex (mPFC) corresponding functionally to human vACC^32–34^, fluorescent miR-151a-5p signal was detected in situ for up to 96 hours post-injection, indicating stable cargo retention in vivo (Supplementary Fig. 2G). To define cell-type specificity of cargo delivery, eNEVs loaded with miR-mimic-TexRed were injected into the mPFC and brains were analyzed 72 hours later (Fig. 3A). Confocal microscopy revealed robust uptake by neurons and microglia, with minimal localization in astrocytes (Fig. 3B).

**Fig. 3:**
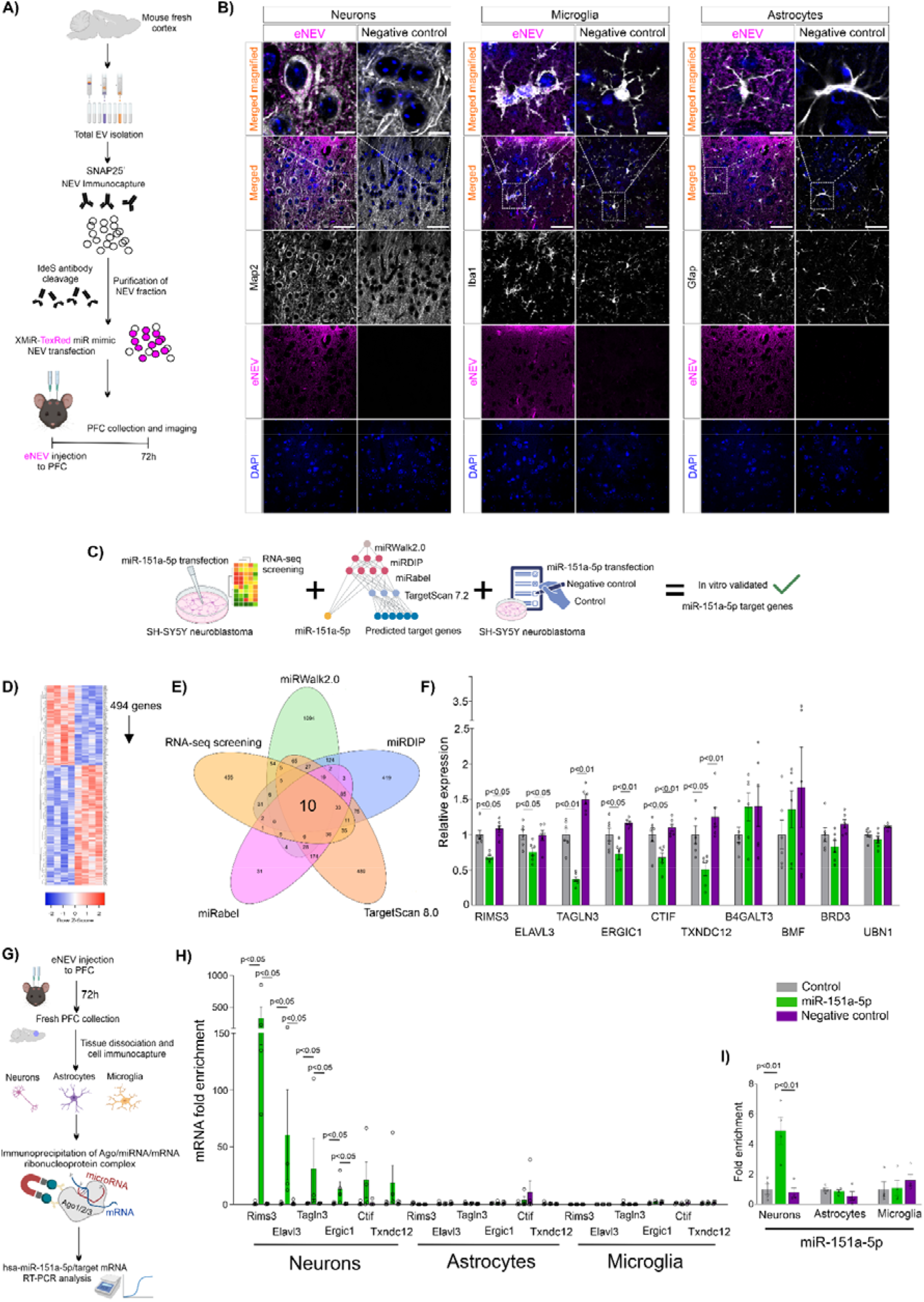
eNEVs enriched with miR-151a-5p selectively deliver cargo to neurons and regulate neuron-enriched genes. A) Schematic of engineered neuron-derived extracellular vesicle (eNEV) production for in vivo cargo-delivery studies. Total EVs were isolated from fresh mouse cortex and enriched for NEVs by immunoprecipitation. NEVs were transfected with XMiR Texas Red-conjugated miRNA mimic oligonucleotide, purified and bilaterally injected into the mouse mPFC; 1.4 μg EV protein equivalent in 1 μl PBS per side). Brains were collected 72 h after injection for histological analysis of fluorescent cargo distribution. B) Representative confocal images of coronal mouse mPFC sections 72 h after eNEV injection (n = 4). Texas Red-labelled miRNA mimic cargo was detected mainly in neuronal cell bodies labelled with Map2 (left panels) and was also detected in Iba1-immunoreactive microglia (middle panels), whereas minimal to no co-localization was observed with Gfap-immunoreactive astrocytes (right panels). Images represent 0.45-μm optical sections through the mPFC. High-magnification scale bar, 10 μm; low-magnification scale bar, 50 μm. DAPI was used to counterstain nuclei. To assess autofluorescence and nonspecific signal, mPFC sections from mice injected with eNEVs carrying non-labelled miR-151a-5p were used as negative controls (n = 1). Rows show, from top to bottom: merged high-magnification images of all channels (DAPI, blue; eNEV cargo, magenta; cell-type marker, grey), merged low-magnification images showing tissue distribution, cell-type marker channel, eNEV cargo channel and DAPI channel. C) Schematic of the combined in vitro and in silico strategy used to identify miR-151a-5p target genes. SH-SY5Y cells were transfected with 50 nM miR-151a-5p, and transcriptomes were profiled by RNA-seq 72 h later and compared with non-transfected controls. Four independent miRNA target-prediction algorithms were then used to identify commonly predicted miR-151a-5p targets. Candidate genes identified by both RNA-seq and in silico prediction were further validated in vitro using an additional negative-control non-targeting miRNA mimic to control for off-target effects. D) Heat map of genes dysregulated by miR-151a-5p transfection in SH-SY5Y neuroblastoma cells relative to non-transfected controls. In total, 494 genes were significantly downregulated following miR-151a-5p transfection (FDR < 0.05). E) Venn diagram integrating RNA-seq results with four target-prediction algorithms, identifying ten high-confidence candidate miR-151a-5p target genes that were predicted by all algorithms and downregulated in vitro. F) In vitro validation of 10 candidate targets in SH-SY5Y neuroblastoma cells. Gene expression was compared across non-transfected controls (Control), cells transfected with a non-targeting miRNA mimic negative control (NC) and cells transfected with miR-151a-5p (miR-151a-5p). Six genes were confirmed as specific miR-151a-5p targets, as they were downregulated by miR-151a-5p but not by the negative-control mimic: RIMS3 (Kruskal–Wallis: H (2) = 12.12, P = 0.0002; post hoc: Control vs miR-151a-5p, P < 0.01; NC vs miR-151a-5p, P < 0.01; Control vs NC, P = 0.13), ELAVL3 (one-way ANOVA: F (2) = 4.495, P = 0.029; post hoc: Control vs miR-151a-5p, P < 0.05; NC vs miR-151a-5p, P < 0.05; Control vs NC, P = 0.32), TAGLN3 (one-way ANOVA: F (2) = 71.20, P = 0.0001; post hoc: Control vs miR-151a-5p, P < 0.01; NC vs miR-151a-5p, P < 0.01; Control vs NC, P < 0.01), ERGIC1 (one-way ANOVA: F (2) = 10.37, P = 0.0015; post hoc: Control vs miR-151a-5p, P < 0.01; NC vs miR-151a-5p, P < 0.01; Control vs NC, P < 0.05), CTIF (one-way ANOVA: F (2) = 8.76, P = 0.003; post hoc: Control vs miR-151a-5p, P < 0.01; NC vs miR-151a-5p, P < 0.01; Control vs NC, P = 0.12) and TXNDC12 (Kruskal–Wallis: H (2) = 11.80, P = 0.0002; post hoc: Control vs miR-151a-5p, P < 0.05; NC vs miR-151a-5p, P < 0.01; Control vs NC, P = 0.10). Four candidates were not confirmed as selectively regulated by miR-151a-5p: B4GALT3 (one-way ANOVA: F (2) = 1.206, P = 0.32), BMF (one-way ANOVA: F (2) = 0.784, P = 0.47), BRD3 (one-way ANOVA: F (2) = 3.75, P = 0.047; post hoc: Control vs miR-151a-5p, P = 0.14; NC vs miR-151a-5p, P < 0.05; Control vs NC, P = 0.14) and UBN1 (one-way ANOVA: F (2) = 6.66, P = 0.0085; post hoc: Control vs miR-151a-5p, P = 0.12; NC vs miR-151a-5p, P < 0.01; Control vs NC, P < 0.05); n = 6 per group. G) Schematic of Ago–miRNA–mRNA complex immunoprecipitation and RT–qPCR analysis of miR-151a-5p and target-gene enrichment in neurons, astrocytes and microglia isolated from fresh mouse mPFC. Mice received mPFC injections of PBS (Control) or eNEVs enriched with miR-151a-5p (miR-151a-5p), and eNEVs enriched with a non-targeting miRNA mimic negative control (NC). After 72 h, fresh mPFC tissue was collected and neurons, astrocytes and microglia were isolated from each group. Cells were homogenized, Ago–miRNA–mRNA complexes were immunoprecipitated, and enrichment of miR-151a-5p and its six in vitro validated target genes was measured by RT– qPCR. H) RT–qPCR analysis of fold enrichment of six miR-151a-5p target mRNAs in Ago complexes isolated from neurons, astrocytes and microglia after eNEV injection to mPFC. In neurons, four target transcripts were significantly enriched in Ago complexes after injection of miR-151a-5p-loaded eNEVs: Rims3 (Kruskal–Wallis: H (2) = 7.538, P = 0.0107; post hoc: Control vs miR-151a-5p, P < 0.05; NC vs miR-151a-5p, P < 0.05; Control vs NC, P = 0.24), Elavl3 (Kruskal–Wallis: H = 7.423, P = 0.0132; post hoc: Control vs miR-151a-5p, P < 0.05; NC vs miR-151a-5p, P < 0.05; Control vs NC, P = 0.29), Tagln3 (Kruskal–Wallis: H = 7.53, P = 0.0107; post hoc: Control vs miR-151a-5p, P < 0.05; NC vs miR-151a-5p, P < 0.05; Control vs NC, P = 0.24) and Ergic1 (Kruskal–Wallis: H = 6.50, P = 0.030; post hoc: Control vs miR-151a-5p, P < 0.05; NC vs miR-151a-5p, P < 0.05; Control vs NC, P = 0.32). Ctif and Txndc12, although validated as miR-151a-5p targets in vitro, were not enriched in neuronal Ago complexes and showed low overall abundance: Ctif (Kruskal–Wallis: H= 3.50, P = 0.19) and Txndc12 (Kruskal–Wallis: H = 3.23, P = 0.21). No enrichment of miR-151a-5p target genes was detected in astrocytic Ago complexes: Rims3 (Kruskal–Wallis: H = 5.93, P = 0.030; post hoc: Control vs miR-151a-5p, P = 0.19; NC vs miR-151a-5p, P = 0.40; Control vs NC, P = 0.50), Elavl3 (Kruskal–Wallis: H = 0.73, P = 0.74), Tagln3 (Kruskal–Wallis: H = 3.5, P = 0.19), Ergic1 (Kruskal–Wallis: H = 0.27, P = 0.89), Ctif (Kruskal–Wallis: H = 0.94, P = 0.84) or Txndc12 (one-way ANOVA: F = 3.23, P = 0.21). No target-gene enrichment was detected in microglial Ago complexes: Rims3, not detected; Elavl3, not detected; Tagln3 (Kruskal–Wallis: H = 2.45, P = 0.50), Ergic1 (one-way ANOVA: F = 1.86, P = 0.21), Ctif (Kruskal–Wallis: H= 3.90, P = 0.14) and Txndc12 (one-way ANOVA: F = 0.49, P = 0.62). n= 4 per group. I) RT–qPCR analysis of miR-151a-5p fold enrichment in Ago complexes isolated from neurons, astrocytes and microglia after mPFC eNEV injection. miR-151a-5p was significantly enriched in neuronal Ago complexes (one-way ANOVA: F = 15.31, P = 0.0013; post hoc: Control vs miR-151a-5p, P < 0.01; NC vs miR-151a-5p, P < 0.01; Control vs NC, P = 0.28), but not in astrocytic Ago complexes (one-way ANOVA: F = 1.50, P = 0.27) or microglial Ago complexes (Kruskal–Wallis: H = 2.90, P = 0.25). n = 4 per group. All post hoc pairwise comparisons shown in this figure were performed using the Benjamini, Krieger and Yekutieli two-stage linear step-up procedure. For all graphs, data are presented as mean ± s.e.m.

To identify direct miR-151a-5p targets, we combined in vitro screening with in silico prediction (Fig. 3C). SH-SY5Y neuroblastoma cells transfected with miR-151a-5p revealed 494 transcripts significantly downregulated relative to controls (Fig. 3D). Cross-referencing these genes with four independent miRNA target-prediction algorithms yielded ten high-confidence candidates (Fig. 3E). We then compared the expression of these candidates across non-transfected cells, cells transfected with miR-151a-5p, and cells transfected with an off-target miRNA mimic negative control (carrying a random sequence), confirming six direct miR-151a-5p targets: RIMS3 (Kruskal–Wallis: H (2) = 12.12, P = 0.0002), ELAVL3 (one-way ANOVA: F (2) = 4.495, P = 0.029), TAGLN3 (one-way ANOVA: F (2) = 71.20, P = 0.0001) , ERGIC1 (one-way ANOVA: F (2) = 10.37, P = 0.0015), CTIF (one-way ANOVA: F (2) = 8.76, P = 0.003), and TXNDC12 (Kruskal–Wallis: H (2) = 11.80, P = 0.0002) (Fig. 3F).

Finally, to test whether eNEV-delivered miR-151a-5p has a biological effect in vivo through the endogenous RNA interference machinery, we isolated neurons, astrocytes and microglia from dissociated mouse brain following eNEV injection. We then immunoprecipitated the Argonaute (Ago)–miRNA–mRNA complexes from each cell population (Fig. 3G). In this assay, functional binding of the miRNA to its target mRNA should result in the relative enrichment of both molecules within Argonaute (Ago)–miRNA–mRNA complexes. These effector complexes mediate gene silencing by repressing translation, promoting target mRNA destabilization, or, in some cases, inducing Ago-mediated cleavage of the target mRNA. Cell-type purity was confirmed using canonical markers (Supplementary Fig. 3A–C). We observed significant enrichment of miR-151a-5p within neuronal Ago complexes (Fold enrichment = 4.9, one-way ANOVA: F = 15.31, P = 0.0013), with no enrichment in astrocytes or microglia (Fig. 3I). Among validated targets, four genes were co-enriched with miR-151a-5p in neuronal Ago complexes, with the known neuron-specific genes RIMS3 (Fold enrichment = 320.5, Kruskal–Wallis: H (2) = 7.538, P = 0.0107) and ELAVL3 (Fold enrichment = 35.6, Kruskal–Wallis: H = 7.423, P = 0.0132) showing the strongest effects (Fig. 3H). Notably, despite efficient uptake of eNEVs by microglia, neither miR-151a-5p nor its targets were enriched in microglial Ago complexes, suggesting vesicular cargo degradation in lysosomes rather than cytoplasmic release. Together, these data demonstrate that NEVs selectively delivered miR-151a-5p to neurons, where it can become functionally incorporated into RNA silencing machinery, supporting a role for NEVs in targeted, biologically active inter-neuronal communication in the brain.

### 4. miR-151a-5p cargo delivered by eNEVs has antidepressant-like effects

To assess whether eNEV miR-151a-5p cargo can mediate antidepressant-like effects, we used the chronic social defeat stress (CSDS) paradigm, a well-established mouse model that induces depressive-like behaviors persisting for weeks after the stress regimen^35–37^. Following 10 days of CSDS, stress-susceptible mice were identified by behavioral screening using the social interaction test done 24h after CSDS (SIT1). These received stereotaxically injected eNEVs loaded with miR-151a-5p (n = 15), eNEVs carrying a non-targeting miRNA (n = 13), naked miR-151a-5p (n = 9), or PBS (sham; n = 9) into the mPFC. Control mice not exposed to CSDS were microinjected with either miR-151a-5p-loaded eNEVs (n = 9) or PBS (n = 9). Behavior was reassessed after a 4-day recovery period post injection (Fig. 4A). Specifically, we evaluated antidepressant-like efficacy of the eNEV administration using a second social interaction test (SIT2), which is a core readout of CSDS-induced social avoidance, together with complementary assays of anxiety-and despair-like behavior, including the light/dark box and tail suspension tests performed 24h after SIT2. Administration of eNEVs robustly reversed CSDS-induced social avoidance behavior (Fig. 4B). Stress-susceptible mice that received eNEVs displayed social interaction scores comparable to control mice not exposed to CSDS, indicating restoration of social behavior. In contrast, in control mice, eNEV injection had no effect on social behavior, confirming that eNEVs do not alter baseline social behavior. (Fig. 4B). Importantly, neither NEVs carrying a non-targeting miRNA, nor naked miR-151a-5p injections improved behavioral outcomes measured by SIT2 in CSDS-exposed mice, demonstrating that both miR-151a-5p and its encapsulation within NEVs were required for effective neuronal delivery and behavioral rescue and that this effect was specific to miR-151a-5p (Fig. 4B). We observed a similar pattern in the tail suspension test: mice susceptible to stress treated with eNEVs also showed a marked reduction in total immobility time relative to untreated CSDS-exposed mice, whereas control treatments produced no effect (Fig. 4C). Immobility time in this test is considered a measure of despair-like behavior. Therefore, the reduced immobility observed in eNEV-treated stress-susceptible mice indicates that the intervention ameliorated stress-related behavioral deficits. In contrast, we detected no differences in the light/dark box test across treatment groups (Fig. 4D), which is a measure of anxiety-like behavior, suggesting that eNEV-mediated delivery of miR-151a-5p selectively ameliorates depressive-like behaviors rather than anxiety-related responses. Together, these findings support NEVs as efficient neuronal delivery vehicles of miR-151a-5p capable of eliciting antidepressant-like effects in vivo. This highlights their role as mediators of inter-neuronal communication and their therapeutic potential for restoring molecular and behavioral homeostasis in depression.

**Fig. 4:**
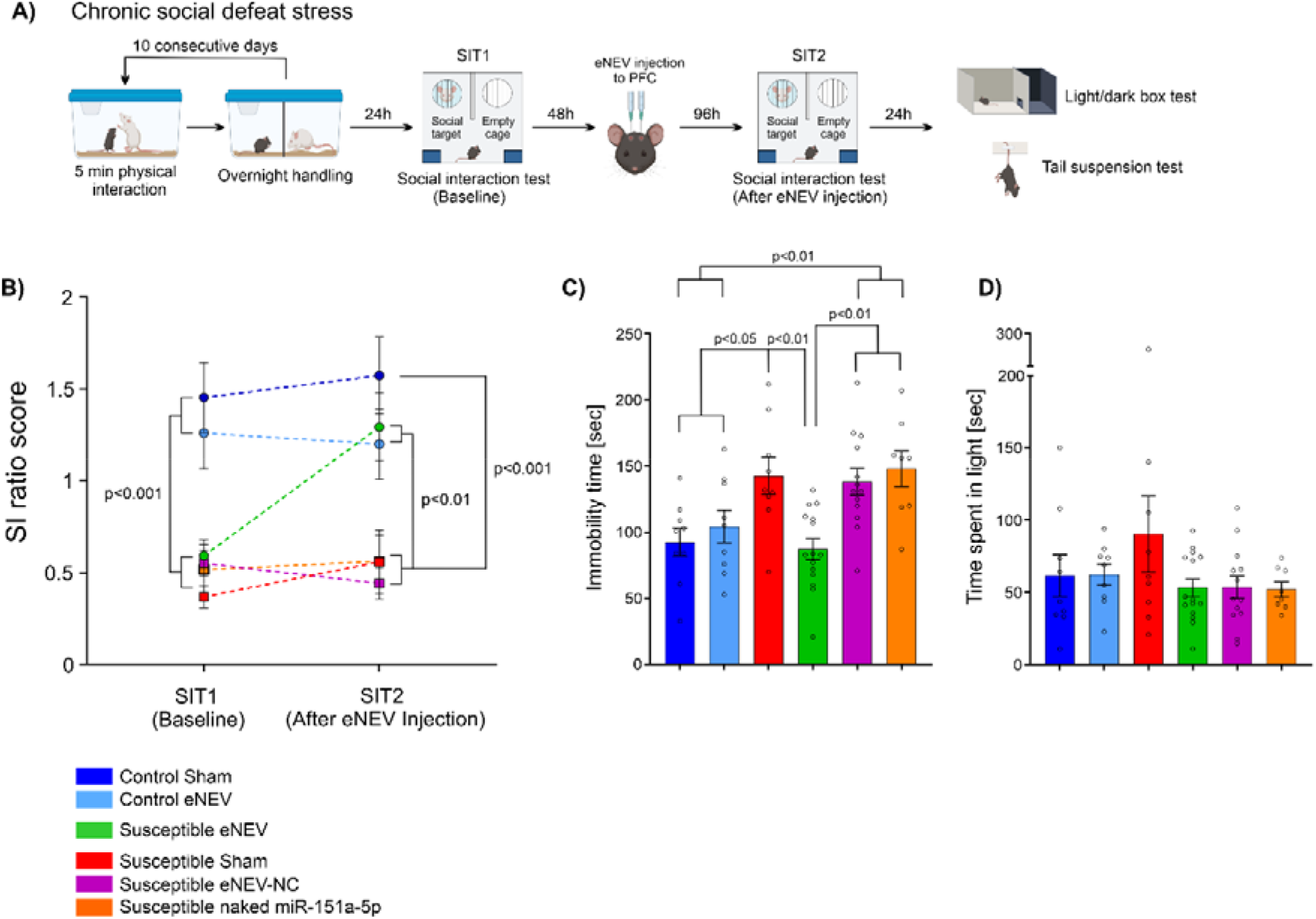
miR-151a-5p cargo delivered by eNEVs has antidepressant-like effects. A) Schematic of the chronic social defeat stress (CSDS) experiment testing the behavioral effects of engineered neuron-derived extracellular vesicles (eNEVs) injected into the mouse mPFC. Twelve-week-old male C57BL/6J mice were subjected to 10 days of CSDS, consisting of 5 min of physical interaction with an aggressive dominant CD-1 mouse followed by overnight sensory exposure in the same cage separated by a perforated transparent Plexiglas divider. At 24 h after the last defeat session, mice were assessed in the social interaction test (SIT1) to measure social avoidance and segregate defeated mice into susceptible and resilient groups. Control and susceptible mice were selected for the intervention phase. Mice then received bilateral mPFC injections of PBS (Control Sham and Susceptible Sham groups), eNEVs loaded with miR-151a-5p (Control eNEV and Susceptible eNEV groups), eNEVs loaded with a non-targeting miRNA mimic negative control (Susceptible eNEV-NC), or non-encapsulated miR-151a-5p (Susceptible naked miR-151a-5p). After 96 h of post-operative recovery, mice were reassessed in the SIT2 and, 24 h later, tested in the light–dark box and tail suspension tests. Control mice in this experiment were not exposed to CSDS paradigm. B) Social interaction ratios measured in stress-susceptible and control mice after CSDS (SIT1) and following eNEV injection into the mPFC (SIT2). Two-way repeated measures ANOVA revealed a significant time × treatment interaction, indicating an effect of miR-151a-5p-loaded eNEVs on CSDS-induced social avoidance (F(5,58) = 2.651, P = 0.031). Post hoc pairwise analysis revealed that at baseline (SIT1), all susceptible groups showed reduced SIT ratios compared with both control non-CSDS control groups (Control Sham and Control eNEV vs all susceptible groups, all P < 0.010), whereas control groups did not differ from each other (Control Sham vs Control eNEV, P = 0.265), and all susceptible groups showed comparable SIT ratios (all pairwise comparisons, P > 0.050). After eNEV injection, at SIT2, only the Susceptible eNEV group showed SIT ratios ≥ 11, reaching values comparable to non-CSDS control groups (Control Sham vs Susceptible eNEV, P = 0.096; Control eNEV vs Susceptible eNEV, P = 0.292). SIT ratios in the Susceptible eNEV group and both control groups were significantly higher than those in Susceptible Sham, Susceptible eNEV-NC and Susceptible naked miR-151a-5p groups (all pairwise comparisons P < 0.010). eNEV injection did not alter basal social behavior in non-CSDS control mice (Control Sham vs Control eNEV at SIT2, P = 0.064). Control Sham, n = 9; Control eNEV, n = 9; Susceptible Sham, n = 9; Susceptible eNEV, n = 15; Susceptible eNEV-NC, n = 13; Susceptible naked miR-151a-5p, n = 9. C) Tail suspension test analysis of behavioral despair, measured as immobility time. Susceptible mice treated with miR-151a-5p-loaded eNEVs showed reduced immobility compared with all other susceptible groups, reaching levels comparable to non-CSDS controls. One-way ANOVA: F (5) = 6.293, P < 0.001; significant post hoc comparisons (P < 0.05) are indicated on the graph. Control Sham, n = 9; Control eNEV, n = 9; Susceptible Sham, n = 9; Susceptible eNEV, n = 15; Susceptible NC, n = 13; Susceptible naked miR-151a-5p, n = 8. D) Light–dark box analysis of anxiety-like behavior, measured as time spent in the light compartment. No significant differences were observed between groups. One-way ANOVA: F (5) = 1.306, P = 0.274. Control Sham, n = 9; Control eNEV, n = 9; Susceptible Sham, n = 9; Susceptible eNEV, n = 15; Susceptible NC, n = 13; Susceptible naked miR-151a-5p, n = 8. All post hoc pairwise comparisons shown in this figure were performed using the Benjamini, Krieger and Yekutieli two-stage linear step-up procedure. For all graphs, data are presented as mean ± s.e.m.

## Discussion

In this study, we established that NEVs act as nanocarriers of molecular signals that change as a function of clinical response to antidepressant treatment. In addition, our results indicate that NEVs can precisely deliver miR-151a-5p cargo to neurons, and elicit molecular changes in the target cell and behavioral changes that mimic those observed with antidepressants. While prior work suggested that EV binding can display cell-type specificity, for example oligodendrocyte-derived EVs preferentially interact with microglia^38^, and neuronal EVs tend to target other neurons^12,13^, direct in vivo evidence that EV cargo becomes functionally integrated into gene regulatory machinery within defined recipient cell types has been limited. Here, we showed that NEVs enriched with miR-151a-5p demonstrated intrinsic neuronal targeting in vivo and, critically, functional incorporation into the neuronal RNA-induced silencing complex (Ago-complex). Although we observed that microglia internalized vesicles, they did not incorporate the cargo into Ago complexes, underscoring that uptake alone does not equate to functional gene regulation and suggesting that cytoplasmic release is tightly controlled and cell-type specific. Consistent with previous observations^39–41^, our observations support a model in which EV-mediated transfer of miRNAs contributes to coordinated regulation of gene expression across neurons, adding a regulatory dimension to classical synaptic signaling.

With eNEVs functioning as neuron-specific nanocarriers, miR-151a-5p mediates the molecular function of recipient neurons once delivered. In our study, miR-151a-5p not only exerted antidepressant-like effects in vivo but was also significantly reduced in both vACC and vACC-derived NEVs from individuals with MDD (Fig. 2G). Interestingly, a study in human post-mortem lateral amygdala which performed a comparative analysis of histone-marked chromatin states using ChromHMM identified a shift from transcriptional to a quiescent state at a putative miR-151a-5p enhancer region (chr8:140789299–140792989, −58 kb)^42^. This transition was observed in individuals with MDD relative to neurotypical controls^42^, suggesting reduced regulatory activity at this locus, which may explain the decreased expression of miR-151a-5p in brains of MDD patients observed in our study. Another exploratory study reported downregulation of miR-151a-5p in total plasma EVs of patients with anxiety^43^. A recent study reported increased levels of miR-151a-3p in total plasma EVs of MDD patients as a function of antidepressant response, accompanied by fMRI changes in ACC^44^, and showed that its global downregulation in the mouse cingulate area 1 (homologous to the human ACC) induces depressive-like behaviors^44^. miR-151a-5p and miR-151a-3p arise from the same precursor but differ in sequence and regulate distinct gene networks. These studies suggest the notion that stress and antidepressant treatment may regulate the miR-151a precursor. However, these effects may occur at different regulatory levels, resulting in arm-specific changes in the abundance of the mature miRNAs.

The target genes of miR-151a-5p that we identified in this study, including RIMS3^45,46^ and ELAVL3^47,48^, implicate its role in presynaptic organization and neuronal RNA stability pathways in relation to MDD and antidepressant response, pointing toward fine-tuning of synaptic homeostasis rather than broad transcriptional repression. This mode of regulation is consistent with the emerging view of miRNAs as stabilizers of molecular networks that buffer synaptic systems against perturbation^49–51^. Within contemporary models of depression that emphasize impaired synaptic plasticity^52,53^ and circuit dysregulation, particularly within medial prefrontal and anterior cingulate networks, reduced miR-151a-5p may reflect destabilization of gene programs necessary for maintaining synaptic equilibrium. Restoration of miR-151a-5p activity during effective antidepressant treatment may therefore represent re-establishment of molecular homeostasis within stress-sensitive circuits rather than a simple, direct pharmacological effect of drug exposure.

Importantly, these molecular effects translate to behavior. In the chronic social defeat stress paradigm known for producing persistent behavioral deficits^35–37^, NEV-mediated delivery of miR-151a-5p reversed social avoidance and despair-like behaviors in stress-susceptible mice, whereas naked miR-151a-5p or NEVs carrying a non-targeting miRNA negative control were ineffective. Notably, the antidepressant-like effects of miR-151a-5p delivered via eNEVs emerged within days following a single administration, in contrast to conventional antidepressants, which typically require repeated dosing to achieve efficacy^54–56^. Such rapid effects are consistent with direct engagement of post-transcriptional regulatory mechanisms, where miR-151a-5p can simultaneously modulate multiple neuronal target genes, bypassing the slower transcriptional and structural adaptations typically required for conventional antidepressant efficacy. These findings suggest that miR-151a-5p is not only a biomarker associated with antidepressant response but also a regulator capable of modulating depressive-like phenotypes when reintroduced into mPFC via EVs. The requirement for vesicular encapsulation underscores that physiological modes of intercellular transfer are critical for effective neuronal engagement. As previously shown, the intercellular specificity of miRNA transfer via EVs is not random but is mediated by specific nucleotide motifs (EXOmotifs) within miRNA sequences that direct their sorting into vesicles^57^. miR-151a-5p contains one of the core characterized EXOmotifs (GGAG)^57^, suggesting that its presence in NEVs is a regulated, rather than stochastic, process. The apparent absence of effects on anxiety-related behavior further suggests circuit-level specificity, consistent with the notion that depressive and anxiety phenotypes are partially dissociable at the level of medial prefrontal circuitry^58,59^.

Several limitations warrant consideration. SNAP25-based isolation captures a heterogeneous population of NEVs derived from multiple neuronal subtypes and brain regions, limiting spatial and cellular resolution. The low abundance of circulating NEVs necessitates relatively large biofluid volumes or highly sensitive detection approaches. The human data remains observational; although the study was longitudinal and pharmacokinetically controlled, causal inference requires experimental manipulation, which is not feasible in humans. eNEVs provide mechanistic insight but do not fully recapitulate endogenous release dynamics, and stereotaxic administration does not model systemic delivery. Furthermore, NEVs carry complex molecular cargo beyond miR-151a-5p, and coordinated effects among multiple regulatory molecules remain to be elucidated. Finally, the intracellular mechanisms governing selective cargo release and engagement in recipient neurons are incompletely understood.

Despite these limitations, this study advances a unified conceptual framework in which SNAP25-positive NEVs function as carriers of neuronal regulatory information and miR-151a-5p emerges as both a biomarker and modulator of synaptic and behavioral plasticity in depression. By integrating longitudinal patient data, brain region-specific postmortem analyses, and in vivo functional experiments, our findings support a model in which depression involves dysregulation not only of neuronal activity patterns but also of intercellular gene regulatory signaling.

## Materials and Methods

### Human post-mortem brain samples

All brain samples used in this study were obtained from the Douglas Brain Bank (RRID:SCR_025991; www.douglasbrainbank.ca). The study was given ethical approval by the Research Ethics Board (REB) of the Centre Intégré Universitaire de Santé et de Services Sociaux (CIUSSS) de l’Ouest-de-l’Île-de-Montréal, and informed consent was obtained from a family member of each individual included in this study. We analyzed three human post-mortem brain regions: vACC, BA4 and LGN obtained from 10 controls (5 females, 5 males) and 20 individuals with MDD (9 females, 11 males). Brain tissue regions were dissected by expert brain bank staff and stored at -80X°C until use. Proxy-based, structured and standardized interviews (SCICI/II) were used as a psychological autopsy tool to obtain clinical information about deceased individuals ^60,61^. Individuals with MDD met the DSM-IV/V criteria for MDD and died by suicide during an episode of major depression. Controls died by natural or accidental causes and did not meet criteria for any major Axis I disorders. Both groups were matched for racial composition, sex proportion, age, pH, PMI, refrigeration delay and RNA integrity number (RIN) of isolated total RNA from all brain regions (Supplementary Fig. 1J). For NEV characterization experiments, we used fresh post-mortem cortical tissue samples (n = 3) from healthy control individuals who died by accidental causes.

### Human CSF samples

CSF samples (n=3) were obtained from healthy control participants^62^ with approval from the Douglas Mental Health University Institute Research Ethics Board. All participants provided written informed consent.

### Human plasma samples

Plasma samples for miRNA profiling and NEV characterization were obtained from healthy controls who took part in the multisite CAN-BIND1 clinical study (NCT01655706)^29,63^. Whole blood samples were collected in EDTA-treated tubes and centrifuged for 10 min at 1500 x g at RT to remove cells from plasma. After centrifugation, plasma supernatants were transferred to new polypropylene tubes, aliquoted and stored at -80 °C until use. The study was approved by Research Ethics Boards at each site and written informed consent was obtained from all participants.

### CAN-BIND-1 clinical study

The study cohort comprised patients with major depressive disorder (n = 142, 89 females and 53 males)) and healthy controls (n = 73, 48 females and 25 males) enrolled in the CAN-BIND-1 study (NCT01655706)^29,63^. Participants aged 18–65 years were recruited between August 2013 and December 2016 across six academic centers in Canada through physician referrals and public advertisements. All participants provided written informed consent, and the study was approved by the institutional ethics boards at each site. Clinical assessments were conducted at baseline and after 8 weeks using the Montgomery–Åsberg Depression Rating Scale (MADRS). Patients met criteria for a current major depressive episode of at least 3 months’ duration and had baseline MADRS scores >24. Exclusion criteria included pregnancy or breastfeeding, primary Axis I diagnoses other than MDD, significant Axis II disorders, substance dependence within the past 6 months, and failure to respond to at least four adequate pharmacological treatments. Healthy controls had no history of Axis I or II disorders, as assessed by the MINI, and had MADRS scores <10. All participants were free of psychotropic medication at baseline. Patients received escitalopram (ESC, 10–20 mg) for 8 weeks, after which treatment response was defined as a ≥50% reduction in MADRS score from baseline. After clinical evaluation of symptoms before and after 8-weeks of treatment we classified subjects to three groups: psychiatrically healthy controls (n=73), responders (n=71) and nonresponders (n=71). The three groups did not differ in sex, ethnical composition, or age (Supplementary Fig. 1A).

### Escitalopram and S-desmethylcitalopram serum concentration measurements

Serum concentrations of ESC and its major metabolite S-desmethylcitalopram (S-DCT) were measured in CAN-BIND1 participants according to a previously published protocol^64^. Briefly, participants reported the timing of their last medication dose at blood collection, and 100 µL of serum was processed by protein precipitation using an acetonitrile:methanol mixture in the presence of deuterated internal standards. Following centrifugation and dilution, samples were analyzed on a triple quadrupole LC–MS/MS system using a short-gradient separation and selected ion monitoring of compound-specific mass transitions. Quantification was performed using multi-point calibration curves (10–1000 ng/mL), with a lower limit of detection of 5 ng/mL, and assay performance was validated using standard quality control criteria for accuracy and precision.

### Isolation of total EV population from human plasma and CSF by size-exclusion chromatography (SEC)

Total populations of plasma and CSF EVs were isolated by using size exclusion chromatography (SEC). Plasma samples were subsequently centrifuged for 10 min at 1500 x g and 10 min at 10 000 x g to remove cell debris. Next, 2 ml of plasma or CSF were added to the Izon chromatography column with a particle size cut-off 70nm (Izon Science, New Zealand), pre-washed with 0.1um filtered PBS, and run according to the manufacturer’s protocol. 2 ml fractions corresponding to a void buffer volume (fractions -3-0), EV-enriched fractions (fractions 1-5) and late fractions containing small particles and soluble proteins (fractions 8-16) were collected in 5ml Eppendorf tubes. SEC fractions were then concentrated to 300 µl using the Amicon Ultra-4 centrifugal filter units with 100kDa molecular weight cut-off (Millipore Sigma, United States).

### Isolation of total EV population from human and mouse brain tissue by SEC

Total population of EVs from fresh and frozen human and mouse brain cortical samples were isolated according to a previously published protocol^65^. Briefly, small brain tissue chunks (∼20mg) were incubated in Hibernate®-E Medium (Thermo Fisher, USA) with 75 U/ml of collagenase type 3 (#CLS-3, Worthington Biochemical Co., USA) at 37°C for a total of 20 min with gentle agitation, with a ratio of 800 µl of digesting solution per 100 mg of brain tissue, with sporadic pipetting using a wide-bore pipette. Digestion was stopped by addition of the Complete Protease Inhibitor and PhosSTOP phosphatase inhibitor cocktail (Millipore Sigma, USA) in PBS to a final concentration 1 ×, on ice. Samples were centrifuged at 100 × g for 5 min at 4°C to remove cells and debris, and the supernatant was subsequently centrifuged at 10,000 × g for 10 min at 4°C. 2 ml of a supernatant containing EVs and proteins was then applied to a pre-equilibrated SEC column (70nm cutoff, Izon Science, New Zealand) and run with 0.1 µm-filtered PBS according to manufacturer’s instructions. 2 ml fractions corresponding to a void buffer volume (fractions -3-0), EV-enriched fractions (fractions 1-5) and fractions containing small particles and soluble proteins (fractions 8-16) were collected to 5ml Eppendorf tubes. SEC fractions were then concentrated to 300 µl using the Amicon Ultra-4 centrifugal filter units with 100kDa molecular weight cut-off (Millipore Sigma, USA).

### Enrichment for neuron-derived extracellular vesicles (NEVs) by immunoprecipitation

0.2 mg of Dynabeads™ MyOne™ Streptavidin C1 magnetic beads (Cat# 65001, Thermo Fisher Scientific, USA) suspended in 20 µl of PBS with 0.01% Tween20 (BioLabs, USA) were coated with 4µg of biotinylated mouse monoclonal anti-human SNAP25 antibody (Cat# MA5-17610; clone: SP12, Thermo Fisher Scientific, USA) according to the manufacturer’s protocol. Then, the bead-antibody complex (90 µl) was added to 300 µl of purified and concentrated plasma or brain EVs and incubated for 2h at room temperature (RT) under rotation. Beads with SNAP25 positive EVs were separated from the mixture with a magnet, washed four times in PBS containing 0.1% BSA and detached from beads by incubation in 0.1 M glycine-HCl (pH 3.0) for 5 min at RT, followed by neutralization with 1 M Tris-HCl (pH 7.4). Eluted NEVs were used for particle sizing and miRNA analyses. A mixture containing the NEV-depleted EV population after immunoprecipitation was frozen and stored at -80 °C for further studies. For protein analysis of NEVs by Western Blot, NEVs were lysed directly on beads with 1x RIPA buffer (Millipore Sigma, USA) supplemented with protease and phosphatase inhibitors, and beads were subsequently removed magnetically.

### EV concentration and size distribution analysis

NEV concentration and size distribution in samples from the CAN-BIND cohort were measured using a Spectradyne nCS1 instrument (Spectradyne, USA). Samples were diluted in 10× PBS to achieve the optimal particle concentration range recommended by the manufacturer, loaded into disposable C-400 cartridges, and analyzed according to the manufacturer’s protocol. Data acquisition and analysis were performed using Spectradyne software. Particle concentration and size distribution were reported for particles ranging from 65–400 nm.

For NEV engineering experiments, particle concentration and size distribution were assessed by nanoparticle tracking analysis (NTA). Samples were diluted in PBS to a final volume of 0.5 ml and analyzed using a NanoSight NS300 instrument (Malvern Panalytical, UK) according to the manufacturer’s instructions. Data were acquired and processed using NanoSight software. All measurements were performed in at least three technical replicates.

### Quantification of EV markers, SNAP25 and albumin across SEC fractions from human brain, CSF and plasma

We isolated total EV populations from plasma, CSF and fresh human brain according to protocols mentioned above and then SEC fractions were concentrated on Amicon Ultra-4 centrifugal filter units with 10kDa molecular weight cut-off (Millipore Sigma, United States) to concentrate the fractions and prevent the loss of soluble proteins. Concentrated SEC fractions were then sonicated for 2 mins. As a reference for this study to show fractions with SNAP25 not associated with EVs, we used a recombinant free SNAP25 protein (Abcam, UK) diluted in PBS at a concentration of 0.1 µg/ml that was processed by SEC similarly to the rest of the samples. ELISAs were then used to measure the concentrations of SNAP25 (Cat# KE00031, Proteintech, USA), CD9 (Cat# LS-F6853, LSBio, USA), CD81 (Cat# EKC33080, Biomatik, Canada), CD63 (Cat# LS-F26529, LSBio, USA) and albumin (Cat# EKU02214, Biomatik, Canada) in all SEC fractions collected from the plasma (n=3), CSF (n=3) and brain (n=3), according to manufacturers’ protocols. Protein concentrations were calculated from calibration curves using a four-parameter logistic (4-PL) regression model. Values below the limit of detection were assigned as zero.

### Western blot

Human and mouse brain tissue samples were homogenized in ice cold 1x RIPA (Millipore Sigma, USA) with protease and phosphatase inhibitor cocktail at a ratio of 1:10 w/v, centrifuged for 15 mins at 12 000 x g and 4°C, and the supernatant containing proteins was collected. For EVs, a suspension of EVs was mixed with ice-cold 10x RIPA with protease and phosphatase inhibitor cocktail (Millipore Sigma, USA) at a ratio of 10:1 and sonicated. Plasma samples were first depleted of albumin according to a published protocol^66^, protein precipitate was washed in acetone and resuspended in 1x RIPA (Millipore Sigma, USA) with protease and phosphatase inhibitor cocktail. Total protein in all samples was measured by Pierce™ BCA Protein Assay according to the manufacturer’s protocol (Thermo Fisher Scientific, USA). 10ug of total protein from EVs, plasma and brain samples were mixed with Laemmli buffer, denatured, resolved by SDS-PAGE and transferred to nitrocellulose membranes by using a Bio-Rad electrophoresis and semi-dry blotting system according to manufacturer’s protocols (Bio-Rad, USA). Membranes with proteins were then subjected to immunoblotting. Membranes were pre-blocked in Tris-buffered saline with 0.1%Tween20 (TBST) containing 3% BSA for 1h at RT with gentle agitation. Next, blocked membranes were incubated overnight at 4°C with primary antibodies targeting human or mouse:

- SNAP25 (#ab109105; Rabbit mAb; 1:500; Abcam, UK),
- NCAM1 (#99746S; XP® Rabbit mAb; 1:1000; Cell Signaling, USA),
- Peripherin (#ab246502; Rabbit mAb; 1:1000; Abcam, UK),
- GFAP (#GA52461-2; Rabbit polyclonal Ab; 1:1000; Agilent, USA),
- ALIX (#MA5-32773; Rabbit mAb; 1:1000; Thermo Fisher Scientific, USA),
- CD9 (#sc-13118; Mouse mAb; 1:500; Santa Cruz Biotechnology, USA),
- APOB (#sc-393636; Mouse mAb; 1:2000; Santa Cruz Biotechnology, USA),
- Calnexin (#ab133615; Rabbit mAb; 1:1000; Abcam, UK),
- Tsg101 (#sc-7964; Mouse mAb; 1:1000; Santa Cruz Biotechnology, USA),
- Bip (#MA5-35606; Rabbit mAb; 1:1000; Thermo Fisher Scientific, USA).
- Vdac (#4661; Rabbit mAb; 1:1000; Cell Signaling, USA).

Next, membranes were washed 3x for 15 mins in TBST and incubated for 1h at RT in TBST with 3% BSA containing corresponding secondary antibodies conjugated with HRP:

- Goat anti-Rabbit IgG (H+L) Cross-Adsorbed Secondary Antibody (#G-21234; 1:10 000; Thermo Fisher Scientific, USA),
- Rabbit anti-Mouse IgG (H+L) Cross-Adsorbed Secondary Antibody (#31452; 1:10 000; Thermo Fisher Scientific, USA).

For Western blots performed on samples from NEV immunoprecipitation we used Clean-Blot™ IP Detection Reagent-HRP (Thermo Fisher Scientific, USA) at a concentration of 1:100 diluted in TBST with 3% BSA to detect primary antibodies without interference from denatured IP antibody fragments that could obscure the signal from proteins of interest. Excess antibody/IP Detection Reagent-HRP was washed off and the blots were incubated for 1 min in Clarity Western ECL Substrate (Bio-Rad, USA) and developed on a ChemiDoc Imaging System (Bio-Rad, USA).

### EV immunogold labeling and transmission electron microscopy (TEM)

We performed immunogold labeling and transmission electron microscopy (TEM) using a previously published protocol^67^ to verify the existence of a neuron-derived EV (NEVs) subpopulation (EVs simultaneously displaying SNAP25 and tetraspanins on their surface) in the total EV population isolated from plasma and brain. In parallel, the morphology of NEVs isolated from human brain and plasma, as well as NEVs and engineered NEVs (eNEVs) from mouse brain, was assessed by TEM without immunogold labeling, following the same protocol^67^. In brief, 5 μl of EV suspension was adsorbed onto glow-discharged 200-mesh carbon-coated copper grids for 30 min at RT. Grids were fixed in 2% paraformaldehyde in PBS (pH 7.4) for 20 min, washed in water (6 × 1 min), and incubated in 50 mM PBS/glycine for 4 × 2 min. Samples were then blocked in 1% BSA in PBS for 5 min and incubated for 1 h with the following primary antibodies:

- SNAP25 (#ab109105; Rabbit mAb; 1:100; Abcam, UK),
- CD9 (#sc-13118; Mouse mAb; 1:500; Santa Cruz Biotechnology, USA),
- CD81 (#sc-166029; Mouse mAb; 1:500; Santa Cruz Biotechnology, USA),
- CD63 (sc-5275; Mouse mAb; 1:500; Santa Cruz Biotechnology, USA).

For isotype controls, EVs were incubated with the corresponding control antibodies under identical conditions:

- Mouse IgG1 kappa Isotype Control (P3.6.2.8.1) (#14-4714-82; 1:500; Thermo Fisher Scientific, USA),
- Rabbit IgG Isotype Control (#02-6102; 1:100; Thermo Fisher Scientific, USA).

Following incubation, grids were washed in 0.1% BSA in PBS (5 × 2 min) and incubated for 1 h with secondary antibodies:

- 12 nm Colloidal Gold AffiniPure® Goat Anti-Rabbit IgG (#111-205-144; 1:20; Jackson Immunoresearch Laboratories, USA),
- 6 nm Colloidal Gold AffiniPure® Goat Anti-Mouse IgG (#115-195-146; 1:20; Jackson Immunoresearch Laboratories, USA).

Nanogold-conjugated secondary antibodies were diluted in 0.1% BSA in PBS. Excess secondary antibodies were removed by washing in PBS (5 × 2 min). Immunogold-labelled EVs were fixed in 1% glutaraldehyde in 100 mM phosphate buffer (pH 7.4) for 5 min, washed in water (6 × 1 min), negatively stained with 2% uranyl acetate for 30 s, and air-dried. All steps were performed at RT. For NEV morphology analysis, the immunogold labelling step was omitted. EVs were processed as above, fixed in 1% glutaraldehyde, washed, negatively stained, and air-dried. Imaging was performed using a FEI Tecnai 12 transmission electron microscope operating at 120 kV.

### BoNT/A protection assay

To determine the membrane topology of SNAP25 in EVs, total EVs isolated from fresh human brain tissue and plasma were subjected to BoNT/A (#4489-ZN-010, R&D Systems, USA) digestion in the presence or absence of Triton X-100 (BioLabs, USA). Three experimental conditions were tested on 20µg of EV protein in 100 µl of PBS:

- (i) untreated EVs, serving as controls;
- (ii) EVs incubated with 2.5 μg BoNT/A and 1% Triton X-100, which permeabilizes EV membranes and allows enzymatic access to both luminal and external SNAP25;
- (iii) EVs incubated with 2.5 μg BoNT/A alone, where intact EV membranes protect luminal SNAP25 while permitting digestion of membrane-exposed protein.

Samples were incubated at 37 °C for 1.5 h and subsequently mixed with 4× Laemmli buffer (3:1 v/v) and heated at 95 °C for 10 min to inactivate the enzyme and denature proteins. Equal sample volumes were analyzed by western blot. SNAP25 levels were quantified relative to Alix, an intraluminal EV marker that is not cleaved by BoNT/A and therefore served as both a loading and experiment specificity control.

### Isolation of total RNA, including miRNAs, from CAN-BIND-1 NEVs

Prior to RNA extraction, NEVs were treated with 20 mg/ml of proteinase K (Thermo Fisher Scientific, USA) for 30 min at 37 °C followed by 10 mg/ml of RNase A (Thermo Fisher Scientific, USA) for 2 min at RT to remove proteins and RNA molecules associated with the external EV surface. Digested NEVs (240 μl) were lysed in 1,200 μl QIAzol reagent, vortexed for 1 min, and incubated for 5 min at RT. Chloroform (240 μl) was then added, samples were vigorously shaken for 15 s, and centrifuged at 12,000 × g for 15 min at 4 °C. The aqueous phase was collected and total RNA, including miRNAs, was purified using the miRNeasy Micro Kit (Qiagen, Germany) according to the manufacturer’s instructions

### Isolation of total RNA, including miRNAs, from brain tissue, cells, EVs and Ago complexes

Human brain tissue was homogenized in QIAzol reagent at a ratio of 10–20 mg tissue per 700 μl QIAzol. Cell samples were homogenized at densities ranging from 5 × 10⁴ to 1 × 10⁶ cells per 700 μl QIAzol. NEVs and engineered NEVs (eNEVs) were lysed in 700 μl QIAzol per 50 μl sample. Ago–miRNA–mRNA immunoprecipitated complexes (10 μl) were dissociated in 700 μl QIAzol. Total RNA, including miRNAs, was isolated using miRNeasy Mini or Micro Kits (Qiagen, Germany) according to the manufacturer’s protocols. RNA concentration and quality were assessed using a NanoDrop 2000 spectrophotometer (Thermo Fisher Scientific, USA) and a TapeStation 4150 automated electrophoresis system (Agilent Technologies, USA).

### Small RNA sequencing and bioinformatic analysis

#### NEV samples

RNA isolated from NEVs was submitted to Norgen Biotek (Ontario, Canada) for library preparation and sequencing. Small RNA libraries were generated using the Norgen Small RNA Library Preparation Kit according to the manufacturer’s protocol and sequenced on an Illumina NextSeq 500 platform using 50-nt single-end reads. Raw sequencing data were processed using CASAVA v1.8+ (Illumina, USA).

#### Leukocyte samples

Whole blood was collected into EDTA tubes and filtered using LeukoLOCK filters (Thermo Fisher Scientific, USA). Total RNA was isolated using a modified LeukoLOCK Total RNA Isolation System protocol incorporating DNase treatment. Libraries were prepared using the NEBNext Small RNA Library Prep Set (New England Biolabs, USA) as previously described^68^. Samples were sequenced at the McGill University and Genome Quebec Innovation Centre (Montreal, Canada) using the Illumina HiSeq4000 with 50nt single-end reads. All sequencing data were processed using CASAVA 1.8+ (Illumina, USA).

#### Data processing and analysis

Sequencing reads were trimmed with cutadapt software to remove adapters and technical sequences. Trimmed, single-end reads were aligned to the human genome (hg38) and miRBase version 22 with no soft-clipping and zero mismatches allowed by using the ERCC’s exceRpt small RNA-Seq pipeline (v.4.6.2) which also quantified aligned miRNAs into counts. Next, counts were normalized according to median expression of the ratios method supplemented in DESeq2 Bioconductor package (release 3.13.). Only miRNAs with at least three read counts in 50% of samples from at least one examined group were kept for statistical analysis. Generalized linear model (GLZ) with negative binomial distribution was used to analyze sequencing data. Extreme values from count data table (Cook’s distance value above 0.1) were filtered out and replaced by predicted value from the GLZ model. Filtered data sets were analyzed by factorial GLZ analysis with interaction of two independent factors: time and response to antidepressant treatment.

### RT-PCR validation of small RNA-seq data

For validation of sequencing results, 3.7 μl of NEV-derived RNA was reverse-transcribed using the TaqMan Advanced miRNA cDNA Synthesis Kit (#A28007, Thermo Fisher Scientific, USA) according to the manufacturer’s protocol with minor modifications. Briefly, miRNAs underwent poly(A) tailing, adaptor ligation, reverse transcription, and 24 cycles of pre-amplification. The resulting cDNA was diluted 1:10 and analyzed in triplicate on a QuantStudio 6 Flex Real-Time PCR System (Thermo Fisher Scientific, USA). Data were acquired using QuantStudio Real-Time PCR Software v1.3. TaqMan Advanced miRNA Assays (#A25576, Thermo Fisher Scientific, USA) specific for hsa-miR-151a-5p (ID: 478505_mir), hsa-miR-320a-3p (ID: 478594_mir) and hsa-let-7b-5p (ID: 478576_mir) were used. Relative expression levels were calculated using the 2^−ΔΔCt method, with hsa-miR-320a-3p serving as endogenous control. Reference miRNA was selected based on expression stability analysis in small RNA-seq data using the NormFinder algorithm^69^.

### miRNA RT–qPCR in EVs, Ago complexes and brain tissue

Equal amounts of total RNA including miRNA were reverse transcribed and analyzed by RT-PCR by using TaqMan microRNA Reverse Transcription kit (#4366597, Thermo Fisher Scientific, USA) and TaqMan microRNA Assays (#4427975, Thermo Fisher Scientific, USA) according to the “TaqMan microRNA assays with custom RT pools and custom preamplification pools protocol” provided by the manufacturer. Reactions were performed in triplicate on a QuantStudio 6 Flex Real-Time PCR System (Thermo Fisher Scientific, USA), and data were collected using QuantStudio Real-Time PCR Software v1.3. Relative miR-151a-5p (ID: 002642) expression was calculated using the 2^−ΔΔCt method. The geometric mean of miR-320a-3p (ID: 002277) and let-7b-5p (ID: 002619) was used for normalization as endogenous controls in EVs and brain tissue. U6 snRNA (ID: 001973) was used as reference gene in Ago IP complexes.

### mRNA RT-PCR

Equal amounts of total RNA were reverse transcribed by using the High Capacity cDNA Reverse Transcription kit (#4368813, Thermo Fisher Scientific, USA) and TaqMan Gene Expression Assays (#4351372, Thermo Fisher Scientific, USA) according to manufacturer’s protocol. RT-PCR reactions of cDNA were run in triplicate using the QuantStudio 6 Flex System. Data was collected by using QuantStudio Real-Time PCR Software v1.3. For specific mRNA detection we used the following TaqMan Gene Expression Assays:

- Human RIMS3 (ID: Hs00207275_m1) Mouse Rims3 (ID: Mm01185520_gH)
- Human ELAVL3 (ID: Hs00154959_m1) MouseElavl3 (ID: Mm00809661_s1)
- Human TAGLN3 (ID: Hs01002328_m1) MouseTagln3 (ID: Mm00451109_m1)
- Human ERGIC1 (ID: Hs00384738_m1) Mouse Ergic1 (ID: Mm00470679_m1)
- Human CTIF (ID: Hs00969548_m1) Mouse Ctif (ID: Mm01255901_m1)
- Human TXNDC12 (ID: Hs00210841_m1) Mouse Txndc12 (Mm00481187_m1)
- Human B4GALT3 (ID: Hs00534104_s1) Mouse B4galt4 (ID: Mm00546324_s1)
- Human BMF (ID: Hs05050813_s1) Mouse Bmf (ID: Mm00506773_m1)
- Human BRD3 (ID: Hs00978980_m1) Mouse Brd3 (ID: Mm00469733_m1)
- Human UBN1 (ID: Hs00950609_gH) MouseUbn1 (ID: Mm01343954_m1)
- Human ACTB (ID: Hs99999903_m1) Mouse Actb (ID: Mm04394036_g1)
- Mouse Gapdh (ID: Mm99999915_g1)

### SH-SY5Y cell culture and hsa-miR-151a-5p transfection

Human SH-SY5Y neuroblastoma cells (ATCC, CRL-2266) were obtained from the American Type Culture Collection and cultured according to the supplier’s recommendations. Cells were maintained in a 1:1 mixture of ATCC-formulated Eagle’s Minimum Essential Medium (ATCC 30-2003) and F-12 medium (Gibco, Thermo Fisher Scientific; Cat# 11765054) supplemented with 5% fetal bovine serum under standard culture conditions (37X°C, 5% CO₂). Culture medium was replaced every 5–7 days, and cells were passaged using trypsin. Low-passage cells (passages 3–5) were used for all experiments. Cells at 60% confluence were transfected with 50nM of mirVana® miR-151a-5p mimic (#4464070, ID: MC11537, Thermo Fisher Scientific, USA) or 50nM of mirVana™ miRNA Mimic, Negative Control (#4464061, Thermo Fisher Scientific, USA) using jetPRIME transfection reagent (Polyplus, France) according to the manufacturer’s instructions. After 72 h, cells were washed with PBS and harvested in QIAzol reagent (Qiagen, Germany) for downstream analyses.

### RNA-seq screening of SH-SY5Y cells transfected with miR-151a-5p

Total RNA extracted from transfected SH-SY5Y cells was used for mRNA library preparation with the Illumina NovaSeq X Plus library preparation kit according to the manufacturer’s protocol. Library preparation and sequencing were performed at the Genome Québec Innovation Centre (Montreal, Canada) on an Illumina NovaSeq X platform, generating paired-end 100-bp reads with an average depth of approximately 50 million reads per sample. Adapter sequences and low-quality bases (Phred score <10) were removed using Cutadapt v2.10, and reads shorter than 20 nucleotides after trimming were discarded. Processed reads were aligned to the human reference genome (hg38) using STAR v2.6.1c with splice junction annotations from Ensembl release 90. Gene-level counts were generated using STAR’s GeneCounts option. Differential gene expression analysis was performed using edgeR v3.34.1. P values were adjusted for multiple testing using the Benjamini–Hochberg procedure, and genes with a false discovery rate (FDR) < 0.05 were considered significantly differentially expressed.

### Target prediction for miRNA cargo of miR-151a-5p

Four target prediction algorithms (miRwalk 2.0, miRDIP, TargetScan 7.2 and miRabel) were used to predict mRNA targets for miR-151a-5p. Targets were only considered if they were identified as a predicted mRNA target on all four algorithms plus were significantly dysregulated in RNA-seq screening, as described in a later section.

### Production of eNEVs

First, total EVs were isolated from fresh cortical tissue collected from 12-week-old C57BL/6J mice (approximately 1.2 g total tissue) according to a protocol described above in the section of the Materials and Methods “Isolation of total EV population from human and mouse brain tissue by SEC”. Next, total EVs were concentrated to 500 µl using Amicon Ultra-4 centrifugal filter units with 100kDa molecular weight cut-off (Millipore Sigma, United States). To enrich NEVs, 500 µl of concentrated EV preparations were incubated for 2 h at RT with 0.5 mg Dynabeads™ MyOne™ Streptavidin C1 magnetic beads (Thermo Fisher Scientific, Cat# 65001) pre-coated with 10 μg biotinylated rabbit monoclonal anti-mouse SNAP25 antibody (Novus Biologicals, Cat# NBP2-81057B; clone SP12) under gentle rotation. Following magnetic separation, beads were washed three times with PBS and resuspended in 100 μl PBS. NEVs were released from the bead–antibody complex using FabRICATOR® (IdeS) enzyme (Genovis AB, Sweden), which specifically and exclusively cleaves IgG antibodies below the hinge region. The enzyme was added at 10 U per 1 μg antibody (100 U total) and incubated for 60 min at 37X°C. After antibody cleavage, magnetic beads were removed, and liberated NEVs were diluted to 500 μl in PBS. The whole procedure was repeated five times, and all NEV preparations were pooled and concentrated to 100 μl using Amicon Ultra-4 centrifugal filter with 100kDa molecular weight cut-off (Millipore Sigma, United States). Total protein concentration was determined using the Pierce™ BCA Protein Assay (Thermo Fisher Scientific, USA). For cargo loading, 25 μg of purified NEVs were transfected with 1 nmol (14 μg) of:

- mirVana™ miR-151a-5p mimic (#4464070, ID: MC11537, Thermo Fisher Scientific, USA),
- miRCURY miR-151a-5p-FITC mimic (#339173, ID: YM00473209-ADB, Qiagen, Germany),
- XMIR Texas Red miR mimic (#XMIR-POS, System Biosciences, USA),
- mirVana™ miRNA Mimic, Negative Control (#4464061, Thermo Fisher Scientific, USA).

Transfection was performed using the Exo-Fect™ siRNA/miRNA Transfection Kit according to the manufacturer’s protocol. Following purification, eNEVs were concentrated to 50 μl using Amicon Ultra-4 centrifugal filters and were used immediately or stored at 4X°C for no longer than 12h.

### Nano-flow cytometric analysis of eNEV transfection efficiency

To assess the efficiency of miR-151a-5p loading into NEVs, eNEVs transfected with FITC-labelled miR-151a-5p were analyzed by nano-flow cytometry. Samples were prepared in 0.1-μm filtered PBS and analyzed at the Centre for Translational Biology, Research Institute of the McGill University Health Centre, using a CytoFLEX Nano flow cytometer (Beckman Coulter, USA) equipped with CytExpert Nano software v1.2. Instrument performance and fluidics were verified daily using CytoFLEX Daily QC Fluorospheres according to the manufacturer’s recommendations. All acquisitions were performed under identical settings: flow rate of 10 μl/min, violet SSC gain of 100, FITC gain of 500, event rates between 2,000 and 6,000 events/sec, abort rates below 3%, and an acquisition time of 5 min per sample. An automated backflush cycle was performed between acquisitions to minimize carryover and cross-contamination. The following controls were included: 0.1-μm filtered PBS alone, free FITC-labelled miR-151a-5p, untransfected NEVs, and Triton X-100-treated eNEVs. Disruption of the vesicle membrane with 1% Triton X-100 was expected to abolish the fluorescent signal if the labelled miRNA was encapsulated within eNEVs. Conversely, the absence of detectable events in samples containing only free miRNA excluded the presence of miRNA aggregates that could mimic vesicle-associated fluorescence. Data were analyzed using CytExpert Nano software and visualized as dot plots.

### Animals

Eight-week-old wild-type male C57BL6/J and CD-1 mice were obtained from Charles River Laboratories Canada and maintained under a 12 h light/12 h dark cycle for the duration of the study. Mice were housed under controlled environmental conditions (22X±X2X°C; 50X±X10% humidity) with unrestricted access to food and water. All behavioral assessments were conducted on male mice of 12 weeks of age during the light phase. Experimental group allocation was blinded to investigators. All procedures involving mice were conducted in accordance with the Canadian Council of Animal Care and approved by the McGill University/Douglas Facility Animal Care Committee.

### Behavioral analyses

Prior to each behavioral assay, animals were habituated to the testing environment for 1X h. All behavioural experiments were performed during the light phase of the light–dark cycle. All apparatus were cleaned thoroughly between sessions to eliminate olfactory cues.

### Chronic social defeat stress paradigm (CSDS)

Chronic social defeat stress (CSDS) was performed as previously described^70,71^. In brief, experimental adult C57BL/6 mice were exposed to 10 consecutive daily defeat sessions using a novel aggressive retired CD-1 breeder mouse screened in advance for reliable aggressive behavior. During each session, the experimental mouse was introduced into an unfamiliar CD-1 mouse compartment for 5Xmin for a direct physical interaction. Following the defeat episode, mice were housed overnight in the adjacent compartment of the same cage as for CD-1 mouse which was divided in two compartments by a transparent and perforated partition that permitted visual, olfactory, and auditory contact between the stressed mouse and the CD-1 aggressor while preventing further physical interaction. Control C57BL/6 mice were housed under identical conditions with a novel littermate daily, without physical contact. All procedures took place between 9:00 AM and 12:00 PM. Subsequent to the last session, animals were individually housed.

### Social interaction test

The social interaction tests were performed according to previously published protocols^70,71^. In brief, 24h after the final CSDS session, and 96h after stereotactic intracranial injection, mice underwent a social interaction test to assess susceptibility to CSDS-induced social avoidance. The test consisted of two consecutive 2.5 Xmin sessions performed in an open-field arena containing a wire-mesh enclosure positioned along one side of the arena. During the first session, the enclosure remained empty, whereas during the second session it contained a novel CD-1 mouse. The time spent within the predefined interaction zone surrounding the enclosure were recorded with Topscan v3 software. Social interaction ratios were calculated as the time spent in the interaction zone in the presence of the social target (CD-1 mouse) divided by the time spent in the same zone in the absence of the target. In SIT1 (screening test) mice with interaction ratiosX<X1 (more time spent away from social target) were classified as stress susceptible and were taken for further experiments, whereas mice with ratiosX≥X1 (more time spent with a social target) were considered as resilient and were not included in the further study. Next, during SIT2 when response to the treatment was measured and mice with ratiosX≥X1 were considered as responders to the treatment, whereas mice with ratiosX<X1 were considered as nonresponders. Results from CSDS stressed mice were compared to non-stressed controls.

### Light–dark box

Anxiety-like behavior was assessed using a light–dark box apparatus consisting of two interconnected 20X×X20Xcm chambers. The lit chamber was illuminated with 350-400 lux light whereas the adjacent dark chamber was enclosed with an opaque black lid with light intensity less than 5 lux. During the 5 min trial, time spent in each compartment, was monitored by camera and quantified with topscan^72^.

### Tail suspension test

Mice were subjected to the tail suspension test as previously described^73^. Animals were individually suspended by the tail using adhesive tape placed approximately 1.5-2 cm from the tip of the tail and secured to a suspension hook positioned above the apparatus floor. Three mice were tested per session in three different compartments of the apparatus. Sessions lasted 6 min and were video-recorded for subsequent offline analysis. During behavioral scoring, total mobility time was quantified throughout the entire session, with mobility defined as active escape-oriented movements involving the body and hind limbs. Mice with increased time spent immobile were considered to be showing greater depressive-like behavior. Minor forelimb movements and passive swinging caused by momentum were not considered mobility.

### Intercerebral injection of eNEVs to mouse mPFC

Stereotaxic injections of eNEVs to the mouse medial mPFC were performed according to already published protocols^70,74^. Briefly, mice were anesthetized with isoflurane (5% induction, 2% maintenance) and secured in a stereotaxic frame to maintain a flat skull position. Following scalp shaving and disinfection, a midline incision was made to expose the skull, and stereotaxic coordinates were determined relative to bregma and lambda using the Allen Brain Atlas. A burr hole was drilled above the target sites, and a glass capillary filled with eNEV suspension and connected to a nanoinjector (Nanoject III, Drummond Scientific, USA) was lowered to the desired coordinates of the mPFC (coordinates: +2mm (A/P), ±0.5mm (M/L), and -2.7 (D/V) relative to Bregma; according to the mouse brain atlas^75^. A total of 1.4 µg of EV protein equivalent containing approx. 120 ng of miR-151a-5p mimic in 1 µl of PBS buffer was injected bilaterally into mouse mPFC. Injection solutions were delivered slowly (1 µl / 8 mins) to minimize tissue damage and reflux. After injection, the capillary was left in place for 3 min before gradual withdrawal. The hole in the skull was sealed with a bone wax and incisions were closed using sutures. Animals received perioperative analgesia, including local lidocaine at the incision site and postoperative systemic analgesics (carprofen 5mg/kg) according to institutional guidelines. Following surgery, mice were placed in a warmed recovery cage and monitored until fully recovered. For time-dependent eNEV signal analysis in the mPFC, mice were euthanized 24h, 48h, 72h and 96h post injection. Postoperative care and general recovery after stereotactical injections to mPFC took place for 4 consecutive days (96 h). 4-day recovery period was chosen in relationship to the administration of eNEVs and analysis of miR cargo signal presence in experiment shown in Supplementary Fig. 2. Behavioral experiments were conducted in four independent mouse cohorts

### Isolation of neurons, astrocytes and microglia from fresh mouse mPFC

For each biological replicate, prefrontal cortices from three mice were pooled. Tissue dissociation was performed according to a published protocol with minor modifications^76^. Briefly, freshly dissected mPFC tissue was sliced into approximately 0.5-mm sections and incubated in pre-warmed Hibernate-A minus Calcium medium (BrainBits LLC, UK) containing 2 mg ml⁻¹ papain (Worthington Biochemical Corporation) for 30 min at 37X°C with gentle agitation. Next slices were transferred to a fresh Hibernate-A medium (Thermo Fisher Scientific, USA) supplemented with B27 (Thermo Fisher Scientific, USA) and GlutaMAX (Thermo Fisher Scientific, USA) and gently triturated using a wide-bore pipette until a single-cell suspension was obtained. The suspension was passed through a pre-wetted 20-μm cell strainer and centrifuged at 300 x g for 5 min at 4X°C. Cellular debris was removed using the Cell Debris Removal Kit (Miltenyi Biotec, USA) according to the manufacturer’s instructions. Purified cells were resuspended in PBS containing 0.5% BSA, divided equally into three aliquots, and subjected to cell-type-specific isolation. Neurons were isolated using the Adult Neuron Isolation Kit (Cat# 130-126-603, Miltenyi Biotec, USA), astrocytes using the Anti-ACSA-2 MicroBead Kit (Cat# 130-097-679, Miltenyi Biotec, USA), and microglia using CD11b MicroBeads (Cat# 130-093-634, Miltenyi Biotec, USA), following the manufacturer’s protocols.

### Argonaute (Ago)–miRNA–mRNA complex Immunoprecipitation

Immunoprecipitation of Argonaute (Ago)–miRNA–mRNA complexes from neurons, astrocytes and microglia isolated from mouse mPFC was performed by using miRNA Target IP kit (Active Motif Inc., USA) according to manufacturer’s protocol. To perform IP, we used approximately 5 x 10^4^ cells per sample and per cell type. miRNA and mRNA enrichment in Ago complexes in each group was determined by RT-PCR and calculations against results from isotypic negative IgG control and transfected control samples according to manufacturer’s instruction.

### Histological analysis of mouse brain and image acquisition

Following completion of the CSDS protocol and eNEV administration mouse brains were dissected from the skull, rinsed in saline, and fixed in 10% neutral-buffered formalin for 48 h at 4X°C. Fixed tissues were washed 2 x 1 h in saline, 1 x 1 h in 1:1 mixture of saline/ethanol and 2 x 1 h in 70% ethanol. Next, brains were dehydrated in increasing concentrations of ethanol and processed using the standard paraffin embedding protocol^77^. Formalin-fixed paraffin-embedded brains were cut into 20 µm coronal sections on a microtome (Leica Biosystems, Germany) and were mounted on SuperFrost Plus™ microscopic slides (Fisher Scientific, USA). Sections were deparaffinized and rehydrated according to a published protocol^78^. Heat-mediated epitope recovery was performed in anhydrous citric acid solution (1.92 g l⁻¹, pH 2.0) for 30 mins at 100X°C. Tissue sections were rinsed briefly in ddH2O following by three 5 min washes in PBS. Washed tissue sections were blocked and permeabilized for 1 h at RT in PBS containing 10% goat serum and 0.05% Triton X-100. Sections were incubated overnight at 4X°C with following primary antibodies:

- Map2 (#ab183830; Rabbit mAb; 1:2000; Abcam, UK),
- Gfap (#ab302644, Goat mAb; 1:1000; Abcam, UK),
- Iba1 (#ab178846; Rabbit mAb; 1:2000; Abcam, UK).

Primary antibodies were diluted in PBS containing 0.1% Tween-20 and 5% BSA. After washing, sections were incubated with the appropriate secondary antibodies:

- Alexa Fluor® 647 AffiniPure® Donkey Anti-Rabbit IgG (#711-605-152; 1:200; Jackson Immunoresearch Laboratories, USA),
- Alexa Fluor® 488 AffiniPure® Donkey Anti-Rabbit IgG (#711-545-152, 1:200; Jackson Immunoresearch Laboratories, USA),
- Alexa Fluor® 488 AffiniPure® Donkey Anti-Goat IgG (#705-545-147; 1:200; Jackson Immunoresearch Laboratories, USA)

Incubation with secondary antibodies lasted 1 h at RT, after which tissue sections were washed in PBS, mounted using VECTASHIELD mounting medium containing DAPI (# H-1800, Vector Laboratories, USA) and stored at 4X°C until imaging. To verify signal specificity, both fluorescently labelled eNEVs carrying FITC- or Texas Red-conjugated miR-mimic and control eNEVs carrying non-fluorescent miR-151a-5p were analyzed.

For time-course experiments assessing eNEV distribution and accumulation, whole-slide images were acquired at 20x magnification using an Olympus VS120 slide scanner (Olympus, Japan) under identical exposure settings across all samples. Images were subsequently analyzed in QuPath v0.3.2.

For cell-type colocalization studies, sections were imaged using an Olympus FV1200 upright confocal microscope equipped with a motorized stage (Olympus, Japan). Images from four mice receiving fluorescent eNEVs and one control mouse receiving non-fluorescent eNEVs were acquired using a ×60 objective (NA = 1.42), with a resolution of 1,600 × 1,600 pixels and a z-step size of 0.45 μm. Laser power, detector gain, acquisition speed, and exposure parameters were kept constant across samples. Imaging settings were optimized to minimize lipofuscin autofluorescence and maximize signal-to-noise ratio.

### The Human Protein Atlas, GTEx and snRNA-seq in silico data exploration

The Genotype-Tissue Expression (GTEx) database of human normal tissue gene expression was used to explore tissue-specific gene expression differences between five candidate markers of neuronal origin: SNAP25, ATP1A3, L1CAM, NCAM1, MAP1B and NRXN3. The data used for the analyses described in this manuscript were obtained from: the <u>GTEx Portal</u> on 05/10/2024. To analyze the expression levels of candidate marker genes in different cell populations from human dlPFC we explored our previously generated single-nucleus RNA-seq database^28^. Raw sequencing data (FASTQ files) for the female dlPFC is available on GEO (accession number: <u>GSE213982</u>) and for the male cohort is available on GEO (accession number: <u>GSE144136</u>). Protein expression scores for all tested targets were obtained from the Human Protein Atlas (https://www.proteinatlas.org/).

## Statistical analysis

Data were assessed for normality using the Shapiro–Wilk test and for homogeneity of variance using the Brown–Forsythe test. Comparisons between two groups were performed using unpaired two-tailed t-tests for normally distributed data or Mann–Whitney tests when normality assumptions were not met. For comparisons involving three or more groups, we used ordinary one-way ANOVA followed by Tukey’s or the Benjamini, Krieger and Yekutieli two-stage linear step-up post hoc tests for normally distributed data, and Kruskal–Wallis followed by the Benjamini, Krieger and Yekutieli two-stage linear step-up post hoc tests for non-normally distributed data. Two-way ANOVA, Two-way repeated measures ANOVA or GLM mixed model followed by Tukey’s or the Benjamini, Krieger and Yekutieli two-stage linear step-up post hoc tests were used to analyze experiments involving two independent factors, including longitudinal analyses over time. Statistical approaches used for small RNA-seq and RNA-seq analyses are described in the corresponding Methods sections. Data are presented as mean ± s.e.m. Statistical significance thresholds are indicated in the corresponding figure legends and graphs, with P-values < 0.05 considered statistically significant. All analyses were performed using Statistica v 13.3 (TIBCO Software) or GraphPad Prism v 9.5.1 (GraphPad Software).

## Acknowledgements

Dariusz Żurawek was financially supported by the Bekker Programme from the Polish National Agency for Academic Exchange (PPN/BEK/2019/1/00287/U/00001).

Alice Morgunova was financially supported by Healthy Brains for Healthy Lives Graduate Student Fellowship; Integrated Program in Neuroscience Internal Award; Postdoctoral Fellowship from the McGill-Douglas Max Planck Institute of Psychiatry International Collaborative Initiative in Adversity and Mental Health, an international partnership funded by the Canada First Research Excellence Fund, awarded to McGill University for the Healthy Brains for Healthy Lives initiative.

Daniel J. Müller was funded by CIHR (FRN: 520164).

Claudio N. Soares is a recipient of research grants from the Ontario Centre for Innovation, OBI, CIHR, Atai Therapeutics, Diamond Therapeutics and Eisai. He serves as a consultant for Bayer, Idorsia, AbCellera, Astellas and Diamond Therapeutics.

Valerie H. Taylor is the Science advisor for Health Canada. She is founder of a Biotech company, Taylored Biotherapeutics. She is supported by grants from CIHR and OBI.

Rudolf Uher is supported by the Canada Research Chairs Program (File Number: 950 - 233141).

Benicio N. Frey holds a Homewood Research Chair in Women’s Mental Health and Depression, is supported by grants from Ontario Brain Institute and Brain Canada, and declares research contracts with Johnson & Johnson and Alcediag, and advisory board engagement with Johnson & Johnson and Eisai.

Gustavo Turecki holds a Tier 1 Canada Research Chair in Major Depressive Disorder and Suicide (CRC-2022-00069) and is supported by grants from the Canadian Institutes of Health Research (CIHR; PJT183903, PJT189993, PJT205924), National Institutes of Health (NIH; R01MH131818), and by the Fonds de recherche du Québec through a Research Centre Grant - https://doi.org/10.69777/5230.

This project has been made possible by the Canada Brain Research Fund (CBRF), an innovative arrangement between the Government of Canada (through Health Canada), and Brain Canada Foundation, the Douglas Hospital Research Centre and the Douglas Foundation, and in part by funding from the Canada First Research Excellence Fund, awarded to McGill University for the Healthy Brains for Healthy Lives initiative, and from the Fonds de recherche du Québec - Santé (FRQS)

The CAN-BIND-1 study (NCT01655706) was supported by a grant the Canadian Biomarker Integration Network in Depression (CAN-BIND), with funding support from the Ontario Brain Institute and the Ontario Research Fund – Research Excellence Program. CAN-BIND is an Integrated Discovery Program carried out in partnership with, and financial support from, the Ontario Brain Institute, an independent non-profit corporation, funded partially by the Ontario government. The opinions, results and conclusions are those of the authors and no endorsement by the Ontario Brain Institute is intended or should be inferred. Additional CAN-BIND-1 study funding was provided by the Canadian Institutes of Health Research (CIHR), and unrestricted donations from, Lundbeck, Bristol-Myers Squibb, and Servier. Funding and/or in-kind support was also provided by the investigators’ universities and academic institutions. All study medications were independently purchased at wholesale market values.

The Genotype-Tissue Expression (GTEx) Project was supported by the Common Fund of the Office of the Director of the National Institutes of Health, and by NCI, NHGRI, NHLBI, NIDA, NIMH, and NINDS.

## Data Availability

All data produced in the present study are available upon reasonable request to the authors.

---

**Supplementary Fig. 1:**
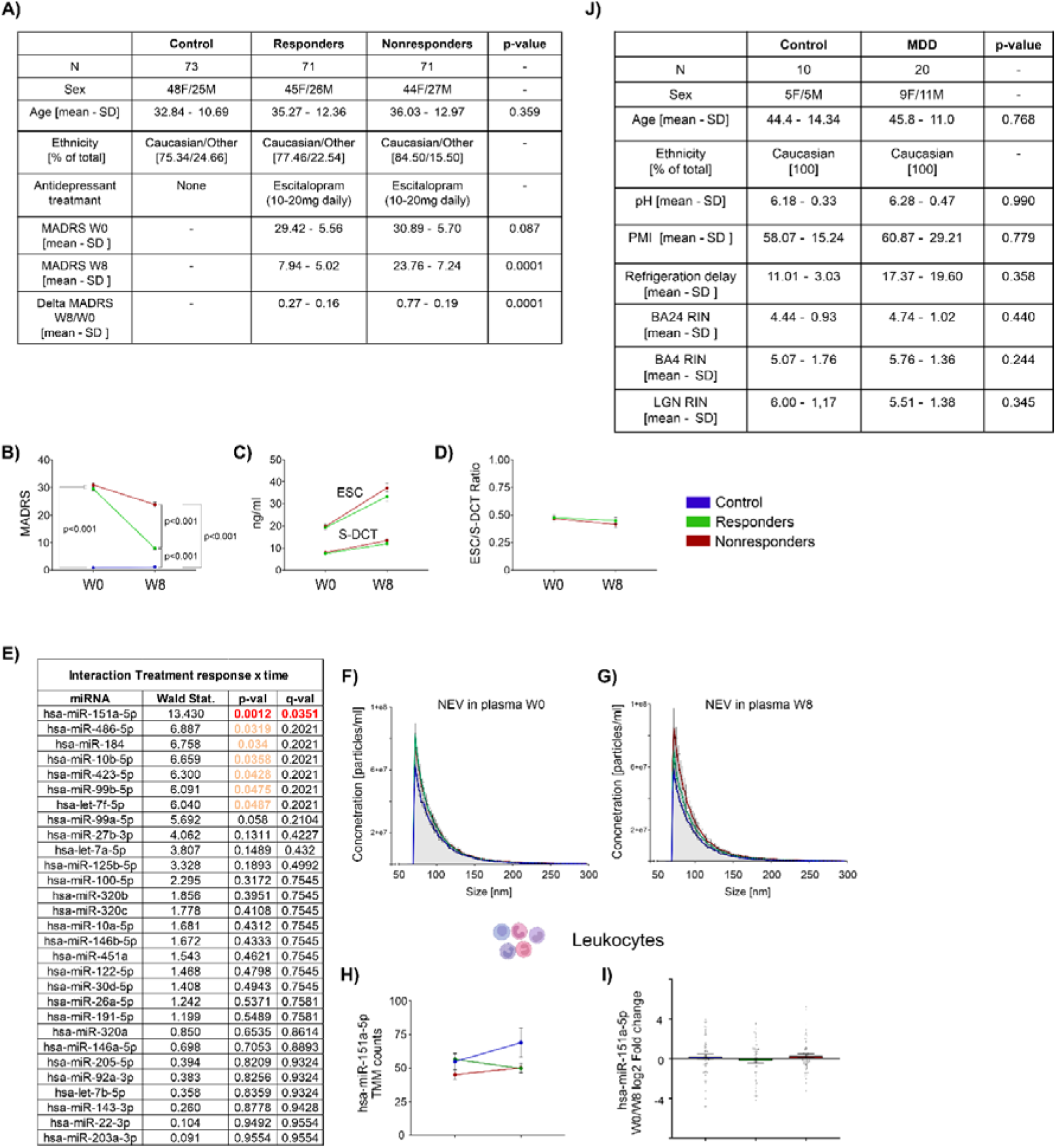
Clinical, pharmacokinetic and technical characterization of the CAN-BIND1 cohort and post-mortem cohort. A) Demographic and clinical characterization of the CAN-BIND1 cohort. Controls, responders and non-responders were balanced for sex distribution and ethnic composition and did not differ in age (Kruskal–Wallis H (2) = 1.856, P = 0.395). B) MADRS scores at baseline (W0) and after 8 weeks of escitalopram treatment (W8) in controls, responders and non-responders. Two-way ANOVA showed a significant time × treatment-response interaction: F(2,212) = 316.9, P < 0.0001. At W0, patients with MDD had significantly higher MADRS scores than controls, with no difference between future responders and non-responders. Tukey’s post hoc tests: W0 controls (n = 73) vs W0 responders (n = 71), P < 0.001; W0 controls vs W0 non-responders (n = 71), P < 0.001; W0 responders vs W0 non-responders, P = 0.187. At W8, MADRS scores were markedly reduced in responders, distinguishing this group from non-responders, in whom symptom reduction was less pronounced. Tukey’s post hoc tests: W8 controls (n = 73) vs W8 responders (n = 71), P < 0.001; W8 controls vs W8 non-responders (n = 71), P < 0.001; W8 responders vs W8 non-responders, P < 0.001. C) Serum concentrations of ESC and its metabolite S-DCT in responders and non-responders, measured two weeks after trial initiation (W0) and two weeks after trial completion (W8). No significant time × treatment-response interaction was observed for ESC (two-way ANOVA: F(1,126) = 1.682, P = 0.197; W0 responders, n = 71; W0 non-responders, n = 70; W8 responders, n = 63; W8 non-responders, n = 65) or for S-DCT (two-way ANOVA: F(1,122) = 1.689, P = 0.196; W0 responders, n = 68; W0 non-responders, n = 70; W8 responders, n = 62; W8 non-responders, n = 64). D) ESC/S-DCT ratio in responders and non-responders, measured two weeks after trial initiation (W0) and two weeks after trial completion (W8). No significant differences in ESC metabolism ratios were observed between groups over time (two-way ANOVA, time × treatment-response interaction: F(1,121) = 0.09516, P = 0.758). E) Summary of all miRNAs detected in plasma NEVs and their statistical analysis. Generalized linear model with negative binomial distribution analysis testing the interaction between time and antidepressant treatment response, followed by FDR correction, identified miR-151a-5p as the only miRNA reaching significance (FDR < 0.05). F) Plasma NEV particle size distribution at W0, expressed as particle concentration (particles/ml). Similar particle distributions were observed across controls, responders and non-responders, with a concentration peak at approximately 100 nm, consistent with EV-sized particles. Mixed-effects GLM testing response group as an independent factor: F (2) = 1.075, P = 0.343. G) Plasma NEV particle size distribution at W8, expressed as particle concentration (particles/ml). Similar particle distributions were observed across controls, responders and non-responders, with a concentration peak at approximately 100 nm. Mixed-effects GLM testing response group as an independent factor: F (2) = 1.886, P = 0.154. H) Small RNA-seq analysis of miR-151a-5p levels in leukocytes, expressed as TMM-normalized counts, at W0 and W8 in controls, responders and non-responders. miR-151a-5p levels did not differ between groups at either time point (two-way ANOVA, time × treatment-response interaction: F(2,396) = 1.569, P = 0.209). W0 controls n = 66; W8 controls, n = 67; W0 responders, n = 66; W8 responders, n = 67; W0 non-responders, n = 69; W8 non-responders, n = 67. I) Log2 fold change in leukocyte miR-151a-5p levels between W0 and W8 in controls, responders and non-responders, measured by small RNA-seq. No significant differences were observed between groups (Kruskal–Wallis: H = 0.836, P = 0.658). Controls, n = 66; responders, n = 64; non-responders, n = 66. J) Demographic and technical characterization of the post-mortem human brain cohort. Control subjects (n = 10) and individuals with MDD (n = 20) did not differ in sex distribution or ethnic composition. Groups also did not differ in age (two-tailed t-test, P = 0.768), brain pH (Mann–Whitney test, P = 0.990), post-mortem interval (PMI; two-tailed t-test, P = 0.779), refrigeration delay (Mann–Whitney test, P = 0.358), or RNA integrity number (RIN) in vACC (two-tailed t-test, P = 0.440), BA4 (two-tailed t-test, P = 0.244) and LGN (two-tailed t-test, P = 0.345). For tables in A and J data are presented as mean ± s.d.; for graphs in B-I, data are presented as mean ± s.e.m.

**Supplementary Fig. 2:**
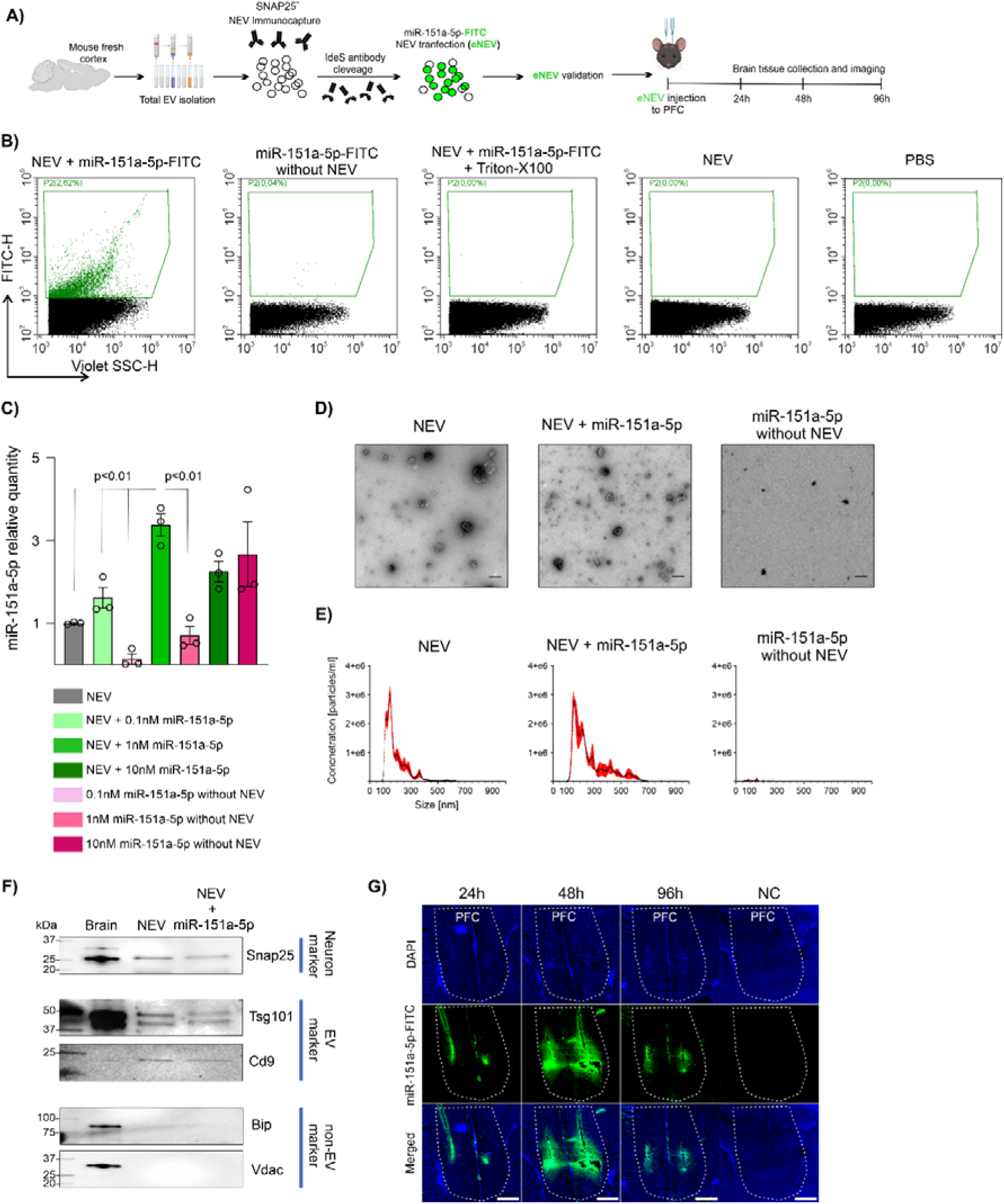
Optimization and characterization of miR-151a-5p-loaded eNEVs for in vivo delivery. A) Schematic of eNEV production for in vivo analysis of cargo distribution in mouse brain. Total EVs were isolated from fresh mouse cortex and enriched for NEVs by immunoprecipitation. NEVs were released from the immunoprecipitation complex by IdeS-mediated digestion of the anti-SNAP25 antibody used for immunocapture, and then transfected with either FITC-conjugated miR-151a-5p or non-labelled miR-151a-5p. Transfection efficiency and the optimal NEV-to-miR-151a-5p ratio were determined using complementary validation approaches. eNEVs loaded with miR-151a-5p–FITC were injected into the mouse mPFC, and brains were collected 24, 48 and 96 h after injection for histological analysis of fluorescent cargo distribution. eNEVs loaded with non-labelled miR-151a-5p were used to control for tissue autofluorescence. B) RT–qPCR analysis of NEV transfection efficiency with miR-151a-5p. Because native brain NEVs contain endogenous miR-151a-5p, untransfected NEVs were used as a reference control. NEVs equivalent to 25 μg protein were transfected with increasing concentrations of miR-151a-5p (0.1, 1 or 10 nM), purified from untransfected miRNA by size-exclusion chromatography, and compared with corresponding free miR-151a-5p plus transfection reagent controls. A ratio of 1 nM miR-151a-5p per 25 μg NEVs provided the best transfection-to-background ratio, with high miR-151a-5p enrichment and minimal contamination by free miRNA aggregates. One-way ANOVA: F(6) = 10.66, P = 0.0002; post hoc comparisons: NEV + 1 nM miR-151a-5p vs untransfected NEVs, NEV + 0.1 nM miR-151a-5p, 0.1 nM miR-151a-5p without NEVs and 1 nM miR-151a-5p without NEVs, all P < 0.01. At the highest ratio, miR-151a-5p levels did not differ between NEV-containing and NEV-free samples (NEV + 10 nM miR-151a-5p vs 10 nM miR-151a-5p without NEVs, P = 0.241), suggesting vesicle saturation and reduced efficiency of SEC-based removal of unincorporated miRNA. n = 3 per group. Post hoc pairwise comparisons were performed using the Benjamini, Krieger and Yekutieli two-stage linear step-up procedure. Data are presented as mean ± s.e.m. C) Representative TEM images of untransfected NEVs, NEVs transfected with miR-151a-5p and miR-151a-5p processed without NEVs. Transfected NEVs retained typical cup-shaped EV morphology, whereas miR-151a-5p processed without NEVs showed no EV-like particles and only minimal aggregate-like material. Scale bar, 100 nm; n = 3 per group. D) NTA analysis of particle size distribution. Both untransfected NEVs and miR-151a-5p-transfected NEVs showed a particle size distribution peaking at approximately 100 nm, consistent with the expected EV size range. No particles of approximately 100 nm were detected in the miR-151a-5p without NEVs control. n = 3 per group. Data are presented as mean ± s.e.m. E) Nano-flow cytometric analysis of miR-151a-5p–FITC loading into NEVs. A distinct fluorescent particle population was detected only in NEVs transfected with miR-151a-5p–FITC, whereas samples containing miR-151a-5p–FITC and transfection reagent without NEVs showed no signal, indicating efficient SEC-mediated removal of free miRNA aggregates. Treatment of miR-151a-5p–FITC-loaded NEVs with 0.1% Triton X-100, used to disrupt EV membranes and release cargo, abolished the fluorescent particle signal, supporting intravesicular localization of the FITC-labelled miRNA. Native NEVs and PBS controls showed no fluorescent events. n = 3 per group. F) Western blot characterization of miR-151a-5p-transfected NEVs. Transfected NEVs retained SNAP25 signal, comparable to native NEVs and brain homogenate, and were positive for the EV markers Tsg101 and Cd9. Both native and transfected NEVs lacked Bip and Vdac, markers of cellular contamination and non-EV material, indicating that miR-151a-5p transfection did not alter NEV biochemical characteristics. n = 3 per group. G) Histological analysis of fluorescent cargo distribution after mPFC injection of miR-151a-5p–FITC-loaded eNEVs. Brains were collected 24, 48 and 96 h after injection and coronal sections were imaged using an Olympus VS120 slide scanner. eNEVs loaded with non-fluorescent miR-151a-5p were used as an autofluorescence control. FITC signal from miR-151a-5p cargo was detectable in the mPFC up to 96 h after injection. Scale bar, 1 mm; n = 1 per group.

**Supplementary Fig. 3:**
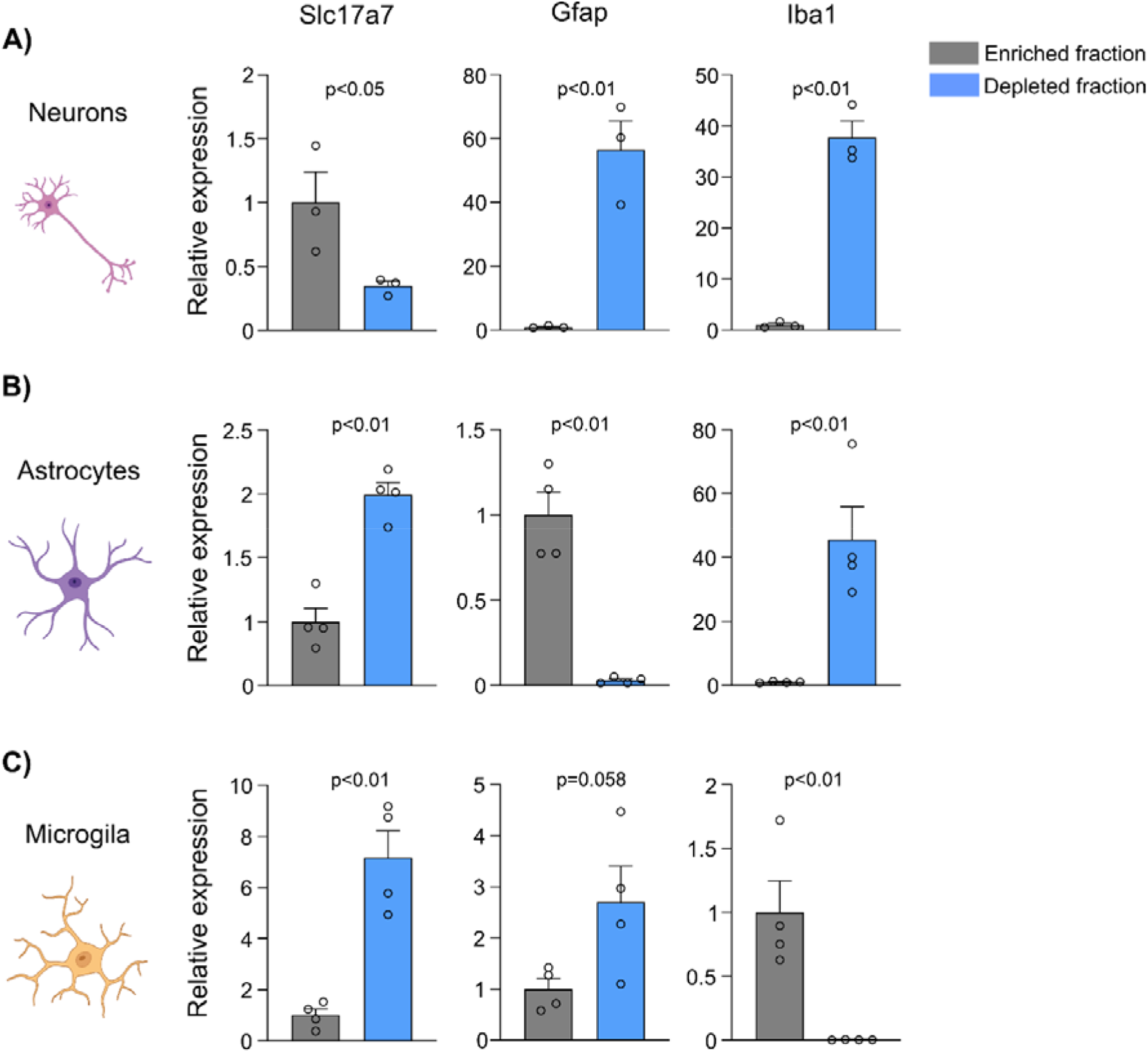
Molecular validation of neuron, astrocyte and microglia isolation specificity from fresh mouse mPFC. A) RT–qPCR analysis of cell-type marker expression in neuron-enriched and neuron-depleted fractions isolated from fresh mouse mPFC. The neuron-enriched fraction showed significant enrichment of the neuronal marker Slc17a7 compared with the neuron-depleted fraction (two-tailed t-test, P < 0.05), whereas the astrocytic marker Gfap and microglial marker Iba1 were significantly depleted (two-tailed t-test, P < 0.01 for both comparisons). n = 3 per group. B) RT–qPCR analysis of marker gene expression in astrocyte-enriched and astrocyte-depleted fractions. The astrocyte-enriched fraction showed significant enrichment of Gfap (two-tailed t-test, P < 0.01) and significant depletion of Slc17a7 and Iba1 compared with the astrocyte-depleted fraction (two-tailed t-test, P < 0.01 for both comparisons), confirming the specificity of astrocyte isolation. C) RT–qPCR analysis of marker gene expression in microglia-enriched and microglia-depleted fractions. The microglia-enriched fraction showed significant enrichment of Iba1 compared with the microglia-depleted fraction (two-tailed t-test, P < 0.01), together with depletion of Slc17a7 (two-tailed t-test, P < 0.01) and a trend toward depletion of Gfap (two-tailed t-test, P = 0.058). n = 4 per group. Marker gene expression was normalized to Gapdh for all cell types. Data are presented as mean ± s.e.m.

## Notes

### Competing Interest Statement

The authors have declared no competing interest.

